# Hip strengthening exercise dosage is not associated with clinical improvements after total hip arthroplasty – a prospective cohort study (the PHETHAS-1 study)

**DOI:** 10.1101/2023.07.12.23292442

**Authors:** Merete Nørgaard Madsen, Lone Ramer Mikkelsen, Michael Skovdal Rathleff, Kristian Thorborg, Thomas Kallemose, Thomas Bandholm

## Abstract

**Purpose:** Postoperative rehabilitation exercise is commonly prescribed after total hip arthroplasty (THA), but its efficacy compared to no or minimal rehabilitation exercise has been questioned. Preliminary efficacy would be indicated if a dose-response relationship exists between performed exercise dose and degree of postoperative recovery. The objective was to evaluate the preliminary efficacy of home-based rehabilitation using elastic band exercise on performance-based function after THA, based on the association between performed exercise dose and change in performance-based function (gait speed) from 3 (start of intervention) to 10 weeks (end of intervention) after surgery.

**Methods:** A pre-registered (NCT03109821) prospective cohort study was conducted. Following primary THA, patients were prescribed home-based rehabilitation exercise using elastic bands. Performed exercise dose (repetitions/week) was objectively measured using attached sensor technology. Primary outcome was change in gait speed (40m fast-paced walk test). Secondary outcomes included patient-reported hip disability. In the primary analysis, a linear regression model was used.

**Results:** Ninety-four patients (39 women) with a median age of 66.5 years performed a median of 339 exercise repetitions/week (1st-3rd quartile: 209-549). Across outcomes, participants significantly improved from 3 to 10-week follow-up. The association between performed exercise dose and change in mean gait speed was 0.01 m/sec [95% CI: -0.01; 0.02] per 100 repetitions.

**Conclusions:** We found no indication of preliminary efficacy of home-based rehabilitation exercise using elastic bands, as no significant and clinically relevant associations between performed exercise dose and changes in outcomes were present. Trials comparing postoperative rehabilitation exercise with no exercise early after THA are warranted.

## Background

### Introduction

Total hip arthroplasty (THA) is commonly performed in patients with severe hip osteoarthritis (OA) to reduce pain and improve function (Gossec et al. 2011), and projections show a significant increase in procedures (Sloan et al. 2018; Pabinger et al. 2018). This challenges health care budgets (Pabinger et al. 2018) and calls for optimised clinical pathways. Functional performance and muscle strength are substantially reduced after THA (Holm et al. 2013; Judd et al. 2014), and postoperative rehabilitation exercise has been recommended (Westby et al. 2014; NICE 2020). However, an evidence-based rehabilitation exercise protocol has not been established (Di Monaco and Castiglioni 2013), and the organization and content of postoperative rehabilitation varies greatly (Smith et al. 2020; Jones et al. 2016; Eulenburg et al. 2015; Feilberg 2016). Using home-based rehabilitation exercise as usual clinical practice is in concordance with clinical guidelines (NICE 2020; Sundhedsstyrelsen 2021) and the findings from a recent systematic review with meta-analysis (Hansen et al. 2019). The systematic review reported, that out-patient rehabilitation exercise with close supervision (minimum two supervised sessions per week) is not superior to home-based rehabilitation exercise with no or very little supervision (a maximum of two supervised sessions after hospital discharge) for both patient-reported and performance-based function, pain and health-related quality of life (Hansen et al. 2019).

Although postoperative rehabilitation exercise in some form is recommended (as opposed to no rehabilitation exercise), the evidence for its effectiveness is inconclusive. Some systematic reviews conclude, that rehabilitation exercise may be superior to no or very little rehabilitation exercise after THA, measured on gait speed and hip abduction muscle-strength (Coulter et al. 2013; Wu et al. 2019), as well as pain and self-reported function (Harris Hip Score) (Wu et al. 2019). Opposed to that, a recent systematic review concluded, that rehabilitation exercise compared to usual care, or no or minimal intervention was not associated with improved patient-reported function or hip muscle strength (Saueressig et al. 2021). A relevant question, which was not addressed in these systematic reviews, is how much exercise patients performed and if the exercise dose is related to the postoperative outcome. Evidence regarding this is sparse, and previous studies have reported conflicting results (Jan et al. 2004; Zech et al. 1. 2015)

The presence of a dose–response gradient is recognized as a criterion for believing in a causal effect (Hill 1965). Therefore, preliminary efficacy would be indicated if a dose-response relationship exists between the amount of performed exercise and degree of postoperative recovery. To investigate a dose-response relationship between post-operative home-based rehabilitation exercise and recovery after THA, objective measures of exercise compliance are needed (Bollen et al. 2014). We have previously validated an in-built sensor attached to an elastic exercise band to monitor compliance to home-based exercise in healthy subjects (Rathleff et al. 2014; Rathleff et al. 2015; Skovdal Rathleff et al. 2013), and started using it for intervention research in clinical populations (Clausen et al. 2018; Husted et al. 2018; Rathleff et al. 2016; Riel et al. 2018). By using this sensor technology, it is possible to objectively quantify performed exercise dose, which improves the validity in studies assessing dose-response relationships and evaluating preliminary efficacy of interventions.

### Objectives

The primary objective was to evaluate the preliminary efficacy of home-based rehabilitation using elastic band exercise on performance-based function after THA, based on the relationship between the performed exercise dose and the change in performance-based function (gait speed measured by 40-m fast-paced walk test) from 3 (start of intervention) to 10 weeks (end of intervention) after surgery (Mikkelsen et al. 2019). The secondary objective was to investigate if a dose-response relationship exists between the performed exercise dose and changes in: hip-related disability, lower-extremity functional performance, and hip muscle strength (Mikkelsen et al. 2019).

## Methods

### Study design and ethics

A pragmatic, single-center, prospective cohort study – “The Pragmatic Home-Based Exercise after Total Hip Arthroplasty – Silkeborg study (PHETHAS-1)” – was conducted. We included patients who were prescribed home-based rehabilitation exercise after THA (usual care) and objectively measured performed exercise dose using sensor technology. Outcomes were measured at week 3 after surgery (start of home-based strengthening exercise, baseline) and at 10 weeks (after 7 weeks of home-based strengthening exercise) after surgery. To minimize sensor-induced influence on exercise compliance and to reduce expectation bias, the participants were informed that the sensor was used to measure how they exercised. They were not told that the focus was on how much exercise was performed nor were they told what the study hypothesis was.

This is the primary study report for PHETHAS-1, which adheres to the STROBE (Strengthening the reporting of observational studies in epidemiology) statement (Vandenbroucke et al. 2007; von Elm et al. 2014). It uses the checklist for cohort studies as well as applicable items from the CONSORT (Consolidated Standards of Reporting Trials) statement (Moher et al. 2012; Schulz et al. 2010) and the REPORT trial guide (Bandholm et al. 2022). Written informed consent was obtained from all participants. The study was reported to The Central Denmark Region Committee on Health Research Ethics and was reviewed as non-notifiable (Inquiry 270/2017). The study was approved by the Danish Data Protection Agency (ref. no: 1-16-02-589-15) and preregistered with ClinicalTrials.gov (Identifier: NCT03109821) (ClinicalTrials.gov.). The full study protocol was published open access, 14 Oct 2019 (Mikkelsen et al. 2019).

### Setting and sampling

The present study was conducted at a public Danish hospital (Elective Surgery Centre, Silkeborg Regional Hospital) that uses the following clinical practice; during admission, patients are instructed to perform unloaded exercises (not part of the intervention studied) at home until their scheduled follow-up visit at the hospital three weeks after surgery. Here, they receive an initial instruction in a home-based rehabilitation exercise program including strengthening exercises to be performed at home without further supervision. Referral to supervised outpatient rehabilitation is initiated in approximately 30% of the patients, based on individual needs. There are no clear-cut criteria for referral to the supervised pathway, but the patient’s preference, rehabilitation goal, functional ability in daily activities, reduced cognitive function and comorbidities are factors influencing the decision.

The study was conducted from 21 April 2017 to 8 January 2020. Exposure was the performed exercise dose during the 7-week intervention period from 3 weeks (baseline) to 10 weeks (follow-up) after THA. A thorough instruction in how to perform home-based strengthening exercise was performed at baseline by physiotherapists from Elective Surgery Centre and represented usual care exercise instruction at our institution. The physiotherapists who did the exercise instruction all had at least 6 months of experience working with THA. Demographics and supplementary descriptive participant variables were collected at baseline (3 weeks after surgery). Performance-based outcome assessments were conducted at baseline and follow-up (10 weeks after surgery) by three physiotherapists who had been thoroughly trained in performing the assessments and who were blinded to exercise compliance data. Patient-reported outcome measures were collected pre-surgery, at baseline and at follow-up (see participant timeline in Appendix A, Table A1, replicated from the published protocol (Mikkelsen et al. 2019)).

Limited availability of equipment (sensors used to measure exercise dose and physical activity) made restricted inclusion necessary and only 2-3 participants could be recruited per week. To mitigate the risk of selection bias, participants were consecutively sampled from random pre-specified assessment programs in the outpatient department. Patients were allocated at random to these assessment programs by a secretary without any influence from personnel involved in the study.

### Intervention

The intervention was the home-based rehabilitation exercise program used in clinical practice at the Elective Surgery Centre, thus, a pragmatic approach was used. In a previously published protocol paper (Mikkelsen et al. 2019), we outlined the intervention in great detail using the exercise-specific Consensus on Exercise Reporting Template (CERT) (Slade et al. 2016) as well as the Template for Intervention Description and Replication (TIDieR) (Hoffmann et al. 2014) – both supplemented with the full set of strength training descriptors as suggested by Toigo and Boutellier (Toigo and Boutellier 2006) (replicated in Appendix A, Table A2). We refer to the published protocol for details (Mikkelsen et al. 2019), but a summarised description is presented below.

All patients received identical instruction in this exercise program, which included strengthening exercises using elastic bands. In short, the intervention under study was initiated immediately after the outcome assessment 3 weeks after surgery (baseline). The strengthening exercises were: hip abduction, flexion and extension with elastic band resistance and sit-to-stands. The prescribed training dosage was two sets with repetitions to contraction failure in each set and a relative load of 10 to 20 repetition maximum (RM), which should be performed every second day (3-4 times a week). Participants were instructed to change the elastic band and obtain a higher load, if they were able to perform more than 20 repetitions in two of three elastic band exercises. Supplemental exercises were daily stretching of hip flexor muscles (stretch 2×30 sec or lying 5-10 min in prone position) and balance exercise (one-legged stance – gradually progressing to 1 min). The prescribed dosage of elastic band exercises was a mean of 630 repetitions per week (range 420 to 840 repetitions per week), but based on previous research (Mikkelsen et al. 2014) and a pilot study conducted prior to this trial (unpublished), a larger variation in actually performed number of repetitions was expected.

The participants were also advised to gradually increase their physical activity level after surgery to comply with the Danish Health and Medicines Authority’s recommendations on physical activity. Furthermore, participants were given instructions on how to handle pain during exercises (reduction of load) (Mikkelsen 2019) and recreational activities. They were advised to contact the hospital in case of increasing exercise-related pain that was not resolved by load reduction, or other complications, such as increase in non-exercise related pain, wound problems or swelling. The pain management guide is available online as extended data for the published protocol (Mikkelsen et al. 2019; Mikkelsen 2019).

### Participants

The inclusion criteria were: age above 18 years, scheduled for primary THA due to OA and ability to understand written and spoken Danish. The exclusion criterion was: referral to supervised rehabilitation in the municipality (instead of the usual care, home-based rehabilitation exercise used in the present study).

### Data collection

#### Demographics and supplementary descriptive variables

Age, gender, height, weight, ASA (The American Society of Anesthesiologists physical status classification system) classification, prosthesis type, prior total joint replacement and length of hospital stay were collected at baseline by the physiotherapist conducting the outcome assessments.

#### Exposure

Performed exercise dose was quantified as the total physiological exercise stimulus (number of repetitions per week) recorded by a sensor (Bandcizer) attached to the elastic exercise band (Rathleff et al. 2014; Rathleff et al. 2015). The sensor automatically switches on, records, and stores exercise data when the elastic exercise band that it is attached to is used. Previously, it has been found valid in measuring date, time of day, number of repetitions, single repetition time-under-tension (TUT), and total TUT during home-based strength training exercises for the lower extremity (Rathleff et al. 2015). Performed exercise dose was also quantified as the number of days per week with strengthening exercises being performed, both based on sensor data and patient-reported data from exercise diaries (see description in Appendix A, Table A3, details on ’mean change in pain after each exercise session’)

#### Outcomes

The primary outcome was the change in gait speed from 3 to 10 weeks after surgery measured by the 40-m fast-paced walk test (Dobson et al. 2013a; Dobson et al. 2013b).

Secondary outcomes were absolute gait speed at 10 week, change in patient-reported function, pain, symptoms and hip-related quality of life measured by Hip disability and Osteoarthritis Outcome Score (HOOS) (Nilsdotter et al. 2003), change in maximal isometric hip flexion and hip abduction strength and change in performance-based lower extremity function measured by the 30-s chair stand test (Dobson et al. 2013a; Dobson et al. 2013b). A detailed description of all study outcomes and measurement tools with supplementary details were presented in the published protocol (Mikkelsen et al. 2019) and are also available in Appendix A, Table A3.

Data collection was continued for participants who stopped exercising but was discontinued if participants explicitly withdrew from the study or if major events or diseases prevented the outcome assessments.

### Data management

Demograhics, supplementary descriptive variables and outcome measurements were entered in EpiData 3.1. Anonymous coding with ID numbers and range checks for data values were used to minimize typing errors. Instead of double entering data as planned, 20% of the participants’s data were validated by two research assistants. Very few and minor errors were found, and further validation was not considered relevant.

Raw Bandcizer data was uploaded to a secure online database. Here, data were accessed, and graphical illustrations of exercise sessions and repetitions were visually inspected. Hereafter performed exercise dose was determined. Due to invalid time-under-tension (TUT) data, and in accordance with the pre-defined contingency plan (Mikkelsen et al. 2019), the exposure variable was changed from TUT to number of repetitions. Reasons are described in detail in the section ‘Deviations from the trial registration and protocol’. The quantification of exercise dose was still challenged, as substantial differences between automatically software-generated and manually-counted number of repetitions were found. To ensure data validity, we therefore manually counted every single repetition for all exercise sets. Also, in case heterogeneity of illustrated repetitions made counting challenging an interpretation level was assigned. Details on this process are available in Appendix A. In one case, data quality was too poor to calculate or count repetitions, and in further two cases, due to sensor failure, no data were obtained. Hence, the latter three cases were not included in the primary analysis (Figure 1).

**Figure 1.**
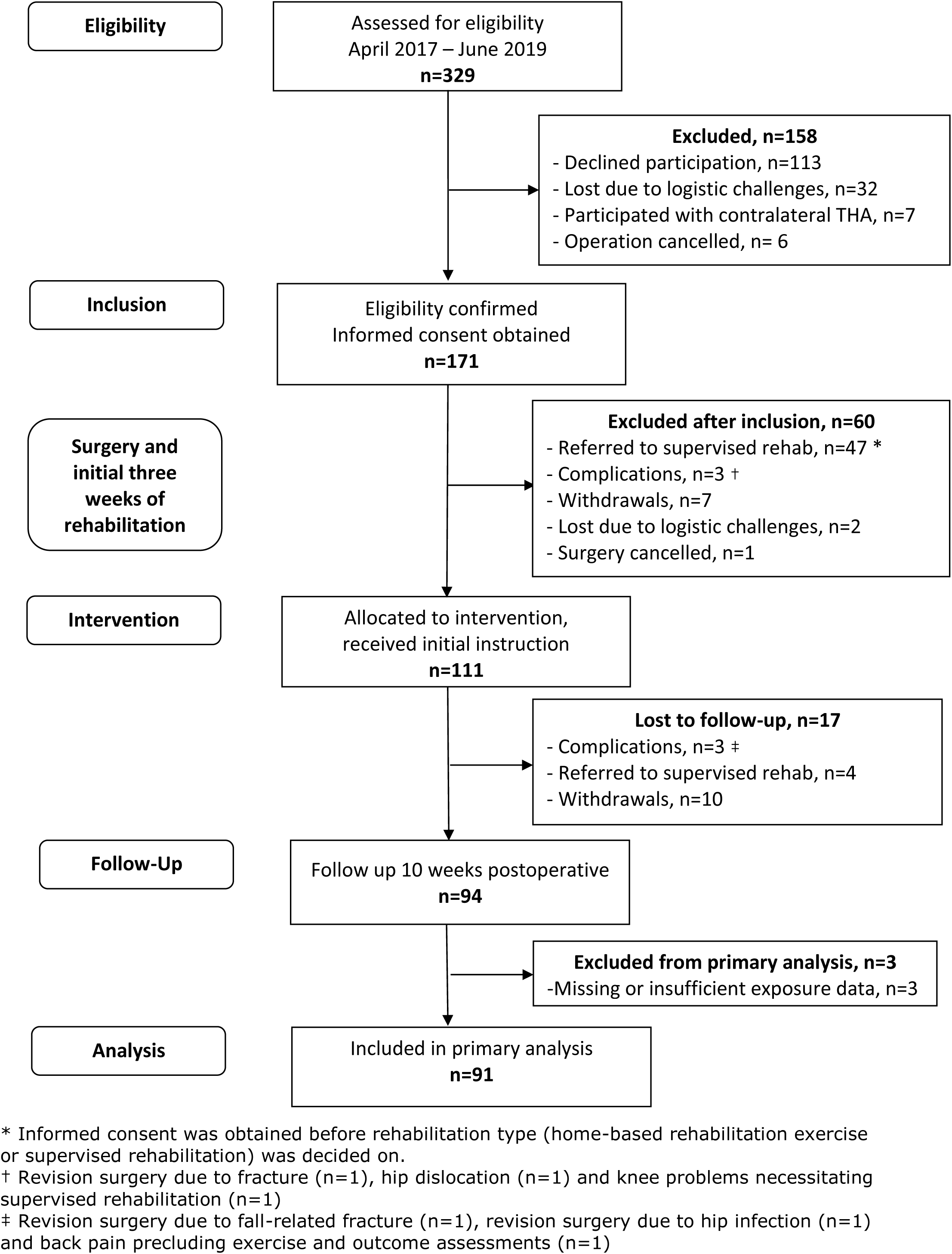
Participant flow (see end of manuscript)

### Sample size

The sample size calculation was outlined in the published protocol, and the procedure is elaborated below. It was based on a linear regression model with exercise dose as a continuous independent parameter and gait speed as the dependent parameter. A minimal clinically important difference (MCID) in the slope for change in gait speed as a function of exercise dose was needed. This MCID slope was based on a previous MCID value for gait speed of 0.2 m/sec (Wright et al. 2011) and the difference between highest and lowest exercise dose, which was estimated to 4 hours (difference in total TUT during the intervention period), based on results from a pilot study conducted prior to this trial (unpublished). Using these values, a MCID slope was represented as a difference in gait speed of 0.2 m/sec from an increase in exercise dose of 4 hours, resulting in a difference of 0.05 m/sec per hour of exercise dose (0.2 m/sec/4 hours). Based on a MCID in slope of 0.05, a standard deviation (SD) for exercise dose of 1.06, a SD for gait speed of 0.16, a power of 90 % and a level of significance of 5%, 88 participants was required. SDs for exercise dose and gait speed were obtained from the previously mentioned pilot study. The sample size calculation was done using the Stata command: sampsi_reg (Mander)

### Statistical methods

A full statistical analysis plan was published as part of the published protocol and formed the basis for the analyses (Mikkelsen et al. 2019). All deviations – with reasons – are provided below. The main analyses are summarized below, while description of exploratory analyses and handling of quantitative continuous and categorical data (e.g. grouping and transformation) are available in Appendix A.

#### Descriptive statistics

Descriptive statistics were performed for demographics, supplementary descriptive variables, pre-surgery HOOS, pre-surgery self-efficacy, exposure, all outcomes (at 3-week, at 10 week and change values) and other pre-specified variables. Categorical variables were presented as frequencies with percentages and continuous variables as means with standard deviation (SD) or medians with 1st and 3rd quartile, depending on data distribution being parametric or not.

#### Primary analysis

The analysis of a dose-response relationship between performed exercise dose and change in gait speed was investigated according to the analysis plan described in the published protocol (Mikkelsen et al. 2019). Based on scatterplots the starting model was a linear regression model with a fixed increase in outcome. R-squared value was low, thus more complex regression models were tested, but without resulting in a model fitting data better. Correlation between change in gait speed and gait speed at baseline was evaluated by scatterplot, and no regression to the mean was indicated. Furthermore, inclusion of the predefined possible confounding variables (self-efficacy at baseline, physical activity during intervention (mean upright time/day and mean number of steps/day), and gait speed at baseline) were evaluated in the model by comparing the dose-response estimates with and without the confounding variables. The normality assumption of the model was evaluated by a quantile-quantile plot and histogram. Estimate of change in gait speed is presented as mean with 95% confidence intervals (CI). Sensitivity analyses of the primary analysis outlined above were performed to test robustness of the estimate. First, outliers in change in gait speed were excluded before estimating the relationship between performed exercise dose and change in gait speed. Secondly, participants, where a high level of interpretation had been used in the count of repetitions, were excluded from the analysis. Thirdly, participants not having used the sensor technology every time or most of the time during exercising were excluded from the analysis. Finally, mean number of exercise days per week (both based on sensor data and data from the exercise diary) was used as performed exercise dose variable in the analysis

#### Secondary analyses

Models similar to the ones used in the primary analysis were used to analyze the dose-response relationship between performed exercise dose and change in patient-reported function measured by the subscale Activity of Daily Living in HOOS (HOOS-adl). The scatterplot of change in HOOS-adl score against performed number of repetitions showed a widespread distribution of data, but linear association was considered the best model for testing association. The scatterplot of change in HOOS-adl against HOOS-adl baseline score indicated regression to the mean, hence, the baseline score was included in the regression model.

Linear regression models were also used to analyze association between gait speed at 10 weeks and performed exercise dose, self-efficacy at baseline, physical activity (mean upright time/day and mean number of steps/day) and gait speed at baseline.

Change in gait speed, HOOS subscales, 30-s chair stand test and maximal isometric hip muscle strength were estimated within each quartile of performed exercise, presented as mean with CI and graphically as boxplots.

### Handling of missing data

As recommended in guidelines, <50% missing items in each subscale of HOOS was accepted (Nilsdotter et al. 2003) and ≤3 missing items in the General Self-efficacy scale was accepted (Schwarzer 2014). For the physical activity data, ≥4 days of data collection with the ActivPAL movement sensor was considered sufficient to calculate mean upright time/day and steps/day (Migueles et al. 2017). In some cases, participants had to stop the performance-based outcome assessments tests due to pain. In these situations, data from the best performance were used no matter if the pre-defined number of test repetitions was met.

In general, we did not use imputation procedures on exposure, but in one case an exception was made. In this case, failure of the BandCizer occurred, leaving the particpant without a sensor for a week before being provided with a new sensor. The participant had perfectly congruence between objectively measured exercise days and self-reported exercise days in the diary, hence, last-observation-carried forward and next-observation carried backwards were imputed to the missing exercise days. As no confounding variables were included in the primary analysis model, no data imputation of possible confounders was performed. Participants lost to follow-up were excluded from the analyses.

### Deviations from the trial registration and published protocol

All predefined analyses have been performed. The exploratory analyses were not pre-defined and registered with ClinicalTrials.gov. However, they were defined in the protocol paper (Mikkelsen et al. 2019), which was published before the end of recruitment and before running any data analyses. Changes to outcomes between registration and protocol publication are reported in the protocol paper (Mikkelsen et al. 2019), hence, only deviations from the published protocol are described in detail below.

First, exposure is presented as number of repetitions per week instead of TUT/week. This change was made according to our pre-defined contingency plan for outcomes (Mikkelsen et al. 2019) and based on the following thorough data assessment. The visual inspection showed a great deal of heterogeneity both within and between exercise sessions and individuals. In general, repetitions seemed to be of shorter duration than recommended and performed with relatively small range of motion. Based on this, we decided to test a sample of exercise sessions, to investigate whether the software’s automatically generated number of repetitions and TUT could be validated against manually-calculated TUT and visually-counted number of repetitions. Based on this test, we realized, that TUT was too imprecise to be considered a valid measure in this study.

Secondly, due to data distribution and data quality, we performed sensitivity analyses for the primary analysis. The sensitivity analyses are described in detail in the section ‘Statistical methods, primary analysis’.

Thirdly, adverse events were grouped as serious adverse events (SAE) and non-serious adverse events. This was decided to provide the reader with the most transparent and clinically relevant overview of data, since several different non-serious events were registered in the category “other”. Classification of SAE was done according to definitions by the U.S Food & Drug Administration (FDA 2016), International Council for Harmonisation of Technical Requirements for Registration of Pharmaceuticals for Human Use (ICH) (ICH 1994) and Ioannidis et al (Ioannidis et al. 2004).

Finally, presentation of summary statistics on body mass index (BMI), pre-surgery HOOS scores, physical activity level and patient-perceived result of surgery were added. Also, supplementary description on pain and exercise data were provided to allow the reader a more detailed insight in data management and exercise compliance.

## Results

### Participants and exposure

Informed consent was obtained from 171 patients, of which 60 were excluded before baseline assessment at intervention start (3 week postoperative). The main reason for exclusion was referral to supervised rehabilitation (n=47). A total of 94 participants completed the study (see Figure 1). Demographics and participant characteristics are presented in Table 1 and Appendix B, Table B1.

**Table 1.** Summary statistics on demographic variables and supplementary descriptive variables*

|  |  |  |
| --- | --- | --- |
| Demographic variables | Age (years)<br>*median (1st; 3rd quartile)* | 66.5 (62; 72) |
|  | Gender<br>*number (percentage)*<br>- Male<br>- Female | 55 (59)<br>39 (41) |
|  | Height (m), n=93<br>*mean (SD)* | 1.75 (0.09) |
|  | Weight (kg), n=93<br>*median (1st; 3rd quartile)* | 81 (72; 95) |
|  | BMI (kg/m<sup>2</sup>), n=93<br>*median (1st; 3rd quartile)* | 26.6 (24.3; 29.4) |
|  | ASA classification, n=92<br>*number (percentage)*<br>- ASA 1<br>- ASA 2<br>- ASA 3 | 26 (28)<br>54 (59)<br>12 (13) |
| Supplementary descriptive variables | Length of hospital stay (days), n=93<br>*number (percentage)*<br>- 0†<br>- 1<br>- 2 | 23 (25)<br>65 (70)<br>5 (5) |
|  | Self-efficacy (mean per answered question)<br>*median (1st; 3rd quartile)*<br>- Pre-surgery, n=89<br>- Baseline (3 weeks), n=88 | 3.3 (2.8;3.7)<br>3.5 (3; 3.8) |
|  | HOOS – pre-surgery<br>*mean (SD)*<br>- ADL<br>- Pain<br>- Symptoms<br>- QOL | 49.3 (15.5)<br>46.5 (14.5)<br>41.3 (16.0)<br>29.7 (13.2) |
|  | Physical activity level, n=81<br>*mean (SD)*<br>- Upright time per day (hours)<br>- Steps per day (numbers) | 5.5 (1.5)<br>6619 (2700) |

\* N=94 unless otherwise stated; † Discharge on the day of surgery
Abbreviations: SD: Standard deviation; BMI: Body mass index; ASA: The American Society of
Anesthesiologists physical status classification system; HOOS: Hip disability and Osteoarthritis Outcome
Score; ADL: Activities of daily living; QOL: Quality of life

The participants performed a median of 2.7 exercise sessions (1st and 3rd quartile: (2.0; 3.2)) and a median of 339 repetitions per week (1st and 3rd quartile: (209; 549)). Hence, compared to the prescribed exercise dose (420-840 repetitions per week) more than 50 % of the participants performed less than the lower limit of recommended number of repetitions per week. Further details on exercise dose are available in Appendix B.

### Primary analysis

Ninety-one participants were included in the analysis. Gait speed improved from a median of 1.45 m/s (1st and 3rd quartile: (0.21; 1.69)) at baseline to 1.74 (1st and 3rd quartile: (1.50; 2.04)) at follow-up, median change: 0.31 (1st and 3rd quartile: (0.21; 0.42), p<0.001). Crude analysis showed a non-significant increase in mean change of gait speed on 0.01 m/s [CI: -0.01; 0.02, p=0.22] per 100 extra repetitions performed per week. Inclusion of the pre-defined possible confounders changed the estimate to values between 0.005 m/s and 0.012 m/s per 100 extra repetitions performed per week. These changes did not change interpretation of the estimate, hence, none of the confounders were included in the analysis model.

### Sensitivity analyses for the primary analysis

Omitting the six outliers in speed change changed the estimate to 0.005 m/s ([CI: -0.005; 0.014], p=0.34) per 100 extra performed repetitions per week, while excluding the three participants where a high degree of interpretation for the estimation of exercise dose was used, changed the estimate to 0.01 ([CI: -0.004; 0.023], p=0.16). Thus, sensitivity analyses changed the estimates, but not to a degree that led to a different interpretation of the results. When using self-reported exercise dose (number of exercise days per week registered in diaries) as exposure, the analysis showed a non-significant mean change in gait speed of 0.03 m/s [CI: -0.01; 0.07], p=0.20) per extra exercise day per week.

### Summary statistics and secondary analyses

Summary statistics for HOOS, 30-s chair stand, and isometric hip muscle strength are presented in Table 2.

**Table 2.** Summary statistics for HOOS, 30-s chair stand and isometric hip muscle strength

| *Outcomes* | *Baseline* | *Follow-up* | *Change* |
| --- | --- | --- | --- |
| *HOOS* |  |  |  |
| *median (1st;3rd quartile)* |  |  |  |
| - ADL | 75 (63; 85)* | 91 (85; 95)* | 13 (6; 21)† |
| - Symptoms | 70 (60; 80)* | 85 (75; 90) | 10 (5; 20)* |
| - Pain | 75 (65; 88)* | 93 (85; 98) | 12 (3; 23)* |
| - QOL | 56 (44; 63)* | 75 (63; 94)* | 19 (6.5; 31)† |
| 30-s chair stand test |  |  |  |
| *median (1st;3rd quartile)* | 12 (10; 15) | 16 (13; 20) | 4 (2; 7) |
| Isometric hip muscle strength (Nm/kg) |  |  |  |
| *mean (95% CI)* |  |  |  |
| - Abduction | 0.76<br>(0.71; 0.82)† | 1.02<br>(0.96; 1.08)† | 0.25<br>(0.21; 0.29)‡ |
| - Flexion | 0.88<br>(0.82; 0.94)* | 1.10<br>(1.03; 1.16)† | 0.22<br>(0.18; 0.26)† |

The table presents values at baseline and follow-up as well as change (baseline to follow-up, 3-10 weeks after surgery) in Hip disability and Osteoarthritis Outcome Score (HOOS), 30-s chair stand and isometric hip muscle strength.
N=94 unless otherwise stated
\* n=93, † n=92, ‡ n=91

A non-significant increase in the HOOS-adl score change of 0.58 ([CI: -0.11;1.26], p=0.10) per 100 extra repetitions performed per week was found. Adjusting for possible confounders resulted in estimates between 0.29 and 0.64 per 100 extra repetitions performed per week. Excluding the most extreme outlier increased change in HOOS-adl to a statistically significant association of 0.75 ([CI: 0.14; 1.36], p=0.02) per 100 extra repetitions performed per week. When excluding all three outliers, the estimated association was 0.67 ([CI: 0.12;1.21], p=0.02).

Change in gait speed, HOOS subscales, 30s chair-stand and maximal isometric hip muscle strength (flexion and abduction) distributed on quartiles of performed exercise dose are presented graphically in Figure 2 and Appendix C, Figures C1-C4. For all outcomes, the exact estimates of change distributed on quartile of performed exercise dose are available in Appendix C, Table C1.

**Figure 2.**
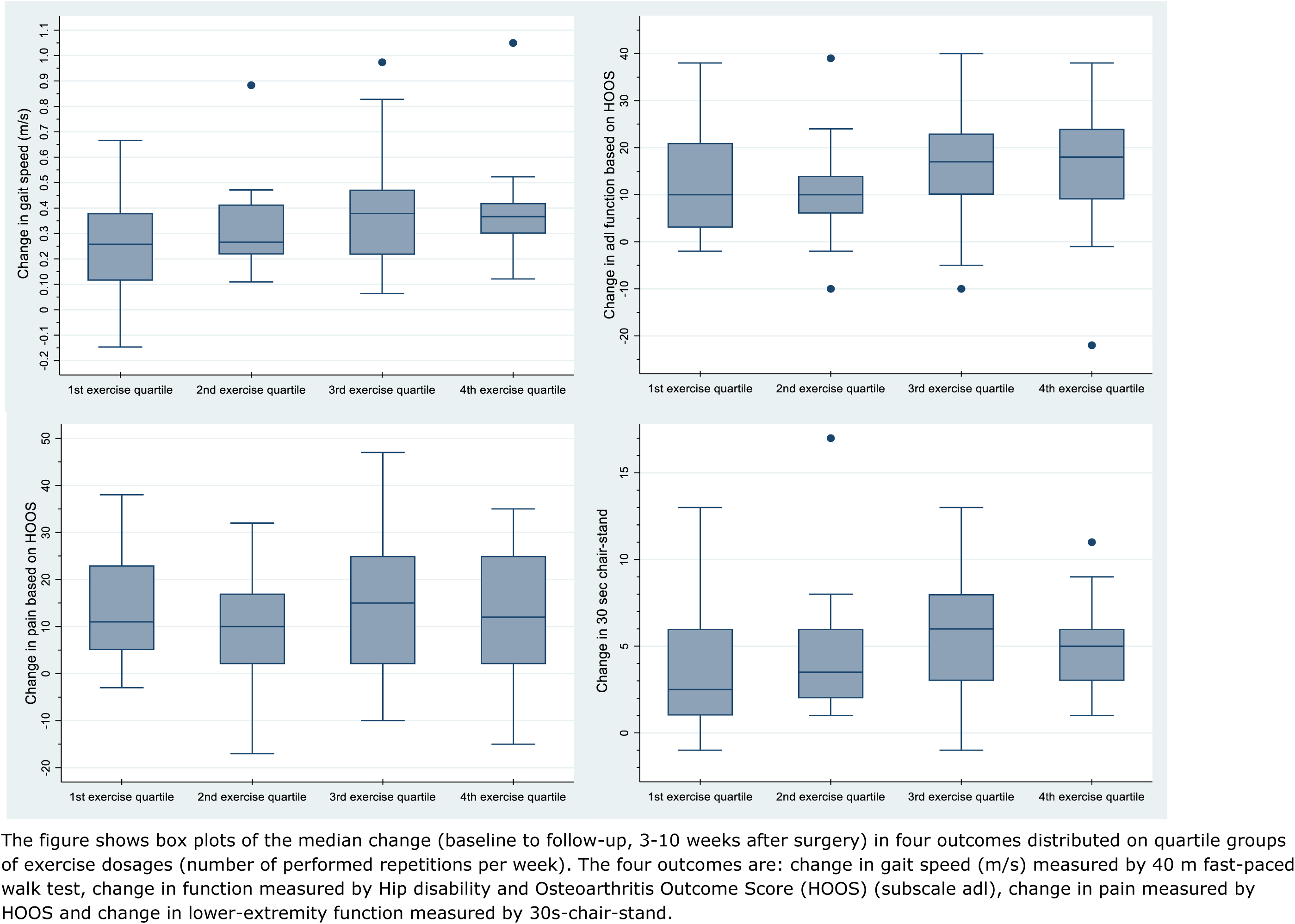
Change in four outcomes distributed on quartile groups of exercise dosages (see end of manuscript)

In the multiple regression analysis of associations between gait speed at 10 weeks follow up and self-efficacy at baseline, 24-hour physical activity, performed exercise dose and gait speed at baseline, small but statistically significant associations were found for the variables: mean upright time/day, mean number of steps/day and gait speed at baseline. Specific results are presented in Appendix C, Table C2.

### Pain and adverse events

Based on each participant’s mean change in pain per exercise session (pain after exercise minus before exercise), the population median change in pain was an increase of 1.5 mm on the visual analogue scale (VAS) (1st and 3rd quartile: (0.2; 3.7)). During intervention period, a total of 57 pain flares (pain change ≥20 mm VAS) in 21 participants occurred following an exercise session. Further details on exercise-related pain are available in Appendix B, Table B5. Among the 94 participants completing the study, two were readmitted due to bleeding and/or spinal headache. In further five participants wound seepage and/or wound infection occurred. Summary statistics on serious and non-serious adverse events are described in detail in Appendix B, Table B6.

### Motivation for and evaluation of exercise, and patient-perceived result of surgery

At baseline, all but one participant were either very motivated or to some degree motivated to perform home-based rehabilitation exercise and 99% of all participants were very or almost certain, that they would comply to the prescribed exercise program. Further summary statistics on items regarding motivation for home-based rehabilitation exercise measured at baseline are available in Appendix B, Table B3.

At follow-up, when evaluating the prescribed exercises, 94 % of the participants were satisfied or very satisfied, and 76% reported that because of participating in the study, they had exercised more than they would have done otherwise. Further summary statistics on items evaluating prescribed exercise are available in Appendix B, Table B4.

The patient-perceived result of surgery at the 10week follow up was rated excellent by 76% of the participants, while 4% reported a fair or poor result. Eighty-eight percent of the participants perceived their hip problem as much better than before surgery, while one participant reported the hip problem to be a little worse. Further details on patient-perceived result are available in Appendix B, Table B4 and Figure B1. The remaining exploratory analyses outlined in Appendix A are presented in Appendix C (Tables C3-7 and Figures C5-7)

## Discussion

### Main findings

No significant linear dose-response association was found between change in gait speed and performed number of repetitions per week. Clinically, the estimated increase in mean change of gait speed on 0.01 m/s [CI: -0.01; 0.02] corresponds to a needed increase of 2000 repetitions per week (1000 reps/week if using the upper limit of the CI) to achieve a minimal clinically important difference of 0.2 m/s (Wright et al. 2011). Hence, the observed associations were not statistically significant, nor were they clinically relevant. Also, no significant linear association was found between the change in HOOS-adl score and the number of performed repetitions per week. However, a sensitivity analysis on HOOS-ADL indicates, that a significant but not clinically meaningful association might be present (0.75 [CI = 0.14; 1.36] per 100 extra repetitions per week). We found no indications of a dose-response relationship when evaluating changes in gait speed, HOOS subscales, 30s chair-stand and isometric maximal hip muscle strength across quartiles of performed exercise. Based on these key findings, preliminary efficacy of home-based rehabilitation exercise was not indicated. No confirmatory conclusions on exercise efficacy can be drawn due to the cohort design without a non-exercise comparator.

### Comparison with previous findings

Two other studies have evaluated associations between exercise dose and clinical improvements in THA populations. Zech et al (Zech et al. 2015) reported that clinical improvements were not associated with the intensity and duration of postoperative exercise therapy in the early phase after THA (Zech et al. 2015), which is in concordance with our findings. Jan et al found contrasting results in their randomized trial comparing participants performing a home exercise program to a control group receiving no exercise (Jan et al. 2004). Participants in the intervention group who exercised more than 50% of the days in the intervention period, achieved greater improvements in muscle strength, gait speed and function, compared to the control group as well as the participants in the intervention group who exercised less than 50% of the days in the intervention period (Jan et al. 2004). The results indicate a dose-response relationship, but the study was conducted more than 1.5 years after surgery (Jan et al. 2004) where spontaneous recovery after surgery likely had no confounding effect. This late timing is in contrast with our focus on early rehabilitation which reflects current clinical practice.

### Explanation of results

The reduced compliance to the prescribed exercises did not seem to affect the overall recovery when compared to outcomes in previous Danish studies reported at similar time-points (Mark-Christensen and Kehlet 2019; Mikkelsen et al. 2014). We have previously shown that patients with THA perceive exercises as a mean to achieve their goals, and that they modify the exercise recommendations according to their needs and individual goals (50). This could be part of the reason why the performed exercise-dose varied substantially among participants. More than 50% of the participants did not perform the number of prescribed repetitions per week, and still more than 75% of the population had an increase in gait speed above the reported clinically meaningful improvement of 0.2 m/s (44). Ninety-one percent of the population reported an excellent or very good patient-perceived result of surgery, 88% rated their hip problem to be much better, and the 10-week scores on HOOS subscales varied between 75 and 93 (100 being the best possible). This may indicate that a dose-response relationship for rehabilitation exercise and post-operative recovery in the early phase after THA does not exist. We speculate that spontaneous recovery determines the recovery trajectory for the most part.

The literature does not provide clear answers as to whether a dose-response relationship between exercise and postoperative outcome should be expected in the THA population. A meta-analysis by Borde et al. investigated resistance training in populations of healthy old adults (Borde et al. 2015) and reported a dose-response relationship between both TUT (per repetition) and training intensity (load) and the effect size for muscle strength (Borde et al. 2015). In addition, a meta-analysis by Ralston et al. reported an association between weekly set volume and strength gain (Ralston et al. 2017). Based on these studies in healthy subjects (Borde et al. 2015; Ralston et al. 2017), a dose-response relationship could have been expected, although exercise responses may differ between healthy adults and adults with severe osteoarthritis recovering from surgery. The American College of Sports Medicine does state that individuals respond differently to resistance training based on training status, past experience and joint health (ACSM 2009) and that a variety of exercise intensities may be effective in the elderly population especially when they start exercising (ACSM 2009). Much like the effect size of spontaneous recovery may blur or exclude an exercise dose-response relationship after THA, the effect size of starting resistance exercise (going from nothing to something) may also blur or exclude an exercise dose-response relationship in previously untrained adults.

### Strengths, limitations and generalizability

A main strength of our study is the use of objectively measured, performed exercise dose. Even though, data quality forced us to use the performed number of repetitions instead of TUT, we still consider the use of performed number of repetitions based on sensor technology to be much more valid than patient-reported data, which can be challenged by non-timely reporting (Stone et al. 2003) and inaccuracy (Prince et al. 2008; Bassett 2003). We are aware, that it could be a limitation, that some interpretation was used in the counting of repetitions, and that 15% of the participants did not use the sensor technology every time or most of the time during exercising. However, interpretation was only used in a minority of cases, and the sensitivity analyses on both issues did not change the interpretation of the primary result. Hence, we do not consider these issues concerning.

A limitation of the study is that about one third of the patients assessed for eligibility declined to participate, inducing a potential risk of selection bias. Possibly, resourceful patients with generally good health are more likely to accept study participation than patients with less resources. Furthermore, 30% of the participants were excluded due to referral to supervised rehabilitation in the municipality. As described in the introduction, there is no clear-cut criteria for this referral, but the excluded participants may be less resourceful, than the group of participants receiving usual care (home-based rehabilitation exercise). Thus, the study results may not be generalized to the less resourceful group of patients receiving a THA.

## Conclusions

We found no indication of preliminary efficacy of home-based rehabilitation exercise using elastic bands, as no significant and clinically relevant associations between performed exercise dose and changes in outcomes were present. Despite no significant association between exercise dose and outcome, participants still improved from baseline to follow-up. Further trials comparing postoperative rehabilitation exercise with no exercise early after THA are warranted.

## Supporting information

ICMJE disclosure forms

## Data Availability

The dataset generated and analysed to provide Figure 1, Table 1 and Appendix B, Table B1 (demographics and participant characteristics) cannot be fully anonymized. Hence, public deposition of these data points is not possible due to Denmarks national legislation (Data Protection Act paragraph 10 and Data Disclosure Proclamation Act) which outline that we can only transfer pseudonymized data to the preprint repository after the Data Protection Authorities approval (Data Protection Act, paragraph 10, section 3, nr. 3.).
Reviewers and others may obtain access to the data (used in the primary and secondary analyses) by reasonable request, on condition that the Danish Data Protection Agency has approved of the data transfer from the Central Jutland Region to the preprint repository. If others are to gain access to the data, the preprint repository shall ensure that there is an adequate legal basis to share the Central Jutland Regions data and ensure that the data is only being processed for scientific research purposes.

## List of abbreviations

ADL: Activities of daily living
ASA: The American Society of Anesthesiologists physical status classification system
BMI: Body mass index
CERT: Specific Consensus on Exercise Reporting Template
CI: 95% confidence intervals
CONSORT: Consolidated Standards of Reporting Trials
HOOS: Hip disability and Osteoarthritis Outcome Score
HOOS-adl: The Subscale Activity of Daily Living in The Hip disability and Osteoarthritis Outcome Score
ICH: International Council for Harmonisation of Technical Requirements for Registration of Pharmaceuticals for Human Use
MCID: Minimal clinically important difference
OA: Osteoarthritis
PHETHAS-1: The Pragmatic Home-Based Exercise after Total Hip Arthroplasty - Silkeborg study
QOL: Quality of life
SAE: Serious adverse events
SD: Standard deviation
STROBE: Strengthening the reporting of observational studies in epidemiology
THA: Total hip arthroplasty
TIDieR: Template for Intervention Description and Replication
TUT: Time-under-tension
VAS: Visual analogue scale

## Declarations

### Ethics approval and consent to participate

Written informed consent was obtained from all participants. The study was reported to The Central Denmark Region Committee on Health Research Ethics and was reviewed as non-notifiable (Inquiry 270/2017). The study was approved by the Danish Data Protection Agency (ref. no: 1-16-02-589-15).

### Consent for publication

Not applicable

### Availability of data and material

The dataset generated and analysed to provide Figure 1, Table 1 and Appendix B, Table B1 (demographics and participant characteristics) cannot be fully anonymized. Hence, public deposition of these data points is not possible due to Denmarks national legislation (Data Protection Act § 10 and Data Disclosure Proclamation Act) which outline that we can only transfer pseudonymized data to the Journal after the Data Protection Authorities approval (Data Protection Act § 10, section 3, nr. 3.).

Reviewers and others may obtain access to the data (used in the primary and secondary analyses) by reasonable request, on condition that the Danish Data Protection Agency has approved of the data transfer from the Central Jutland Region to the Journal. If others are to gain access to the data, the Journal shall ensure that there is an adequate legal basis to share the Central Jutland Regions data and ensure that the data is only being processed for scientific research purposes.

### Authors’ contributions

Conception of the study: LRM, MSR, KT and TB; Design and methodology: All authors Project administration: MNM and LRM; Data acquisition: MNM; Analysis and interpretation of the data: All authors; Statistical expertise: TK; Drafting of the article: MNM; Critical revision of the article for important intellectual content: All authors.

All authors read and approved the final manuscript. Guarantors: Mrs. Madsen and Prof. Bandholm takes responsibility for the integrity of the work as a whole.

## Acknowledgements

The authors would like to thank physiotherapist, Rikke Dahl Nyholm and research assistant Trine Astrup Bech Vestergaard for their invaluable dedication, expertise and help throughout the research process. We would also like to thank the staff and chairs at The Elective Surgery Centre, Silkeborg Regional Hospital for supporting the project, with a special thanks to the involved research assistants, secretaries, nurses and physiotherapists, who’s input and engagement made this study possible. In addition, we would like to extend our sincere gratitude to all the patients who generously participated and gave their time and effort during this study. Lastly, we would like to thank the funding parties, for providing the necessary financial support.

