## Appendix A for "Hip strengthening exercise dosage is not associated with clinical improvements after total hip arthroplasty – a prospective cohort study (the PHETHAS-1 study)"

#### Participant timeline and strength training descriptors

**Table A1.** Participant timeline (replicated from published protocol (1))

| | ***Study period*** | | | |
|---|---|---|---|---|
| Time point | Admission | Baseline | Intervention | Follow up |
| | Pre or post surgery | 3 week visit at the hospital | Week 3-10 post THA | 10 week visit at the hospital |
| **Enrollment** | | | | |
| Eligibility screen | X (pre) | | | |
| Informed consent | X (pre) | | | |
| **Interventions** | | | | |
| Unloaded exercise | X (post) → | | | |
| Strengthening exercise | | Exercise instruction | X | |
| **Assessments** | | | | |
| Performed exercise dose (Elastic band sensor, BandCizer) | | | X | |
| 40m fast-paced walk test | | X | | X |
| HOOS* | X (pre) | X | | X |
| 30s chair stand test | | X | | X |
| Hip muscle strength | | X | | X |
| Pain: VAS** at rest before + after exercise (diary) | | | X | |
| Patient-reported performed exercises (diary) | | | X | |
| Self-efficacy | X (pre) | X | | |
| Physical activity (movement-sensor, ActivPal) | | | X (7 days data collection) | |
| Adverse events | | X | | X |

| | | | | |
|---|---|---|---|---|
| Motivation to exercise as prescribed | | X | | |
| Evaluation of prescribed exercises | | | | X |
| Change in hip problems | | | | X |
| Perception of result after surgery | | | | X |
| Demographics and descriptive variables | | X | | |

* HOOS: Hip disability and Osteoarthritis Outcome Score
** VAS: Visual Analogue Scale

**Table A2.** Strength training descriptors (replicated from published protocol (1))

| | Hip abduction | Hip flexion | Hip extension | Sit-to-stand |
|---|---|---|---|---|
| Load | 15 RM*, acceptable interval: 10-20 RM | 15 RM, acceptable interval: 10-20 RM | 15 RM, acceptable interval: 10-20 RM | 15 RM, acceptable interval: 10-20 RM |
| Repetitions | 10-20 | 10-20 | 10-20 | 10-20 |
| Set per session | Week 1: 1 set (both legs)<br>Week 2-7: 2 sets (both legs) | Week 1: 1 set (both legs)<br>Week 2-7: 2 sets (both legs) | Week 1: 1 set (both legs)<br>Week 2-7: 2 sets (both legs) | Week 1: 1 set<br>Week 2-7: 2 sets |
| Rest between sets | Active rest while exercising opposite leg | Active rest while exercising opposite leg | Active rest while exercising opposite leg | 1-3 minutes |
| Sessions per week | 3-4 (every second day) | 3-4 (every second day) | 3-4 (every second day) | 3-4 (every second day) |
| Duration of training period | 7 weeks | 7 weeks | 7 weeks | 7 weeks |
| Contraction modes | 1 seconds concentric, 1 second isometric, 2 seconds eccentric | 1 seconds concentric, 1 second isometric, 2 seconds eccentric | 1 seconds concentric, 1 second isometric, 2 seconds eccentric | 1 seconds concentric, 1 second isometric, 2 seconds eccentric |
| Rest between repetitions | 0 sec, possible load relieve with one step between reps if needed | 0 sec, possible load relieve with one step between reps if needed | 0 sec, possible load relieve with one step between reps if needed | 0 sec |
| Time under tension | 150 sec / exercise / session at 15 RM | 150 sec / exercise / session at 15 RM | 150 sec / exercise / session at 15 RM | 150 sec / exercise / session at 15 RM |

| | | | | |
|---|---|---|---|---|
| Contraction failure in each set | Yes. The exercise is progressed (elastic band with higher load) when >20 repetitions are accomplished. | Yes. The exercise is progressed (elastic band with higher load) when >20 repetitions are accomplished. | Yes. The exercise is progressed (elastic band with higher load) when >20 repetitions are accomplished. | Yes. The exercise is progressed (backpack with weights) when >20 repetitions are accomplished. |
| Range of motion | Maximum possible | Maximum possible | Maximum possible | Approximately from 90 to 0 degrees of hip and knee flexion. |
| Rest between sessions | 48 hours | 48 hours | 48 hours | 48 hours |
| Anatomical definition of the exercises | Hip abduction is performed in upright standing position with the elastic band looped around both ankles and support by e.g. a solid table. The hip is abducted as much as possible with the toes pointing directly forward and keeping the trunk in upright position. | Hip flexion is performed in upright standing position with the elastic band under the foot of the stance leg and around the ankle of the target leg and support by e.g. a solid table. The target leg is elevated against resistance in a combined hip and knee flexion while keeping the trunk in upright position. | Hip extension is performed in upright standing position with the elastic band looped around both ankles and support by e.g. a solid table. The hip is extended with the ankle flexed to avoid floor contact while keeping the trunk in upright position. | The exercise is performed from standing with equal load on both legs and toes pointing forward. With arms crossed the participants slowly sits down until the chair is just touched and then rises again. |

* RM: Repetition Maximum

**Table A3.** Study outcomes and measurement tools with supplementary details.

| **Outcomes** | **Measurement tool** | **Details** |
|---|---|---|
| Primary | | |
| Change in gait speed from 3 to 10 weeks after surgery | 40-m fast-paced walk test (2, 3). | The 40-m fast-paced walk test was used, as it measures performance-based function and is recommended by Osteoarthritis Research Society International (OARSI) as part of the core set of tests to assess physical function in people with hip or knee OA (2, 3). Also, in a population with hip OA, a high intertester reliability (intraclass correlation coefficient (ICC) 0.95) has been reported (4). Furthermore, patients undergoing THA surgery have reported walking ability to be the most important function to improve (5) |
| Secondary | | |
| Gait speed at 10 weeks | As for primary outcome | As for primary outcome |
| Change in patient-reported function from 3 to 10 weeks after surgery | HOOS, subscale Activities of Daily Living (ADL) (6) | HOOS is a disease-specific patient-reported outcome measure comprising the subscales: symptoms, pain, ADL, function in sport and recreation and hip-related quality of life (6). HOOS is scored on a 0-100 worst to best scale (6). A systematic review has shown HOOS to be valid, reliable (ICC >0.78) and responsible, when evaluating patients undergoing THA (7). |
| Change in patient-reported symptoms from 3 to 10 weeks after surgery | HOOS, subscale symptoms (6) | |
| Change in patient-reported pain from 3 to 10 weeks after surgery | HOOS, subscale pain (6) | |
| Change in patient-reported hip related quality of life from 3 to 10 weeks after surgery | HOOS, subscale hip-related quality of life (6) | |
| Change in lower extremity function from 3 to 10 weeks after surgery | 30-s chair stand test (2, 3). | Assessment was performed using a previously published standardized test procedure (2) where acceptable absolute and relative inter-rater reliability (SEM 7% and ICC 0.88) have been shown after THA (8). |
| Change from 3 to 10 weeks after surgery in maximal | Hand-held dynamometer Power Track II | Maximal isometric hip muscle strength (flexion and abduction) were assessed using standardized test procedures according to previously published methods (9) where |

| | | |
|---|---|---|
| isometric hip abductor muscle strength in the operated leg | Commander in a standardized test procedure (9) | acceptable absolute and relative inter-rater reliability (SEM 7% and 10%; ICC 0.83 and 0.93) have been shown after THA (8). |
| Change from 3 to 10 weeks after surgery in maximal isometric hip flexor muscle strength in the operated leg. | | |
| Other pre-specified variables | | |
| Self-efficacy measured pre-surgery and at 3 weeks after surgery | General self-efficacy scale (10). | The General Self-Efficacy Scale is a validated questionnaire assessing optimistic self-beliefs to cope with a variety of difficult demands in life. Each of the 10 items in the questionnaire is scored on a scale from 1-4 points, with 4 representing the highest level of self-efficacy (10). |
| 24-hour physical activity (mean upright time/day and mean number of steps/day) in week 4 after surgery | ActivPAL movement-sensor | The movement-sensor ActivPAL measures physical activity in terms of time spent in three categories: sitting/lying position, standing and walking. It has been validated in studies in healthy adults (11) and in older adults with a hip fracture (12, 13). The sensor was applied at baseline and used the following week (7 days of data collection). |
| Adverse events | A self-developed chart | Adverse events were registered by the physiotherapist at 3 and 10 weeks after surgery in the pre-defined categories: Hip dislocation, infection, fracture, wound seepage, acute myocardial infarction, deep venous thrombosis, readmission and other |
| Mean change in pain after each exercise session, calculated after 10 week follow-up | Visual analogue scale (VAS) in exercise diary | Pain at rest before and after exercise were registered by the participants in an exercise diary developed for purpose of the present study. Here, participants also registered whether they had exercised, and which exercises they had performed.<br>Data were summarized for each participant as a mean change in pain per exercise session for the entire intervention period. |
| Number of pain flares after exercise sessions calculated at 5 weeks and 10 weeks | | Pain at rest before and after exercise were registered by the participants in an exercise diary developed for purpose of the present study. An increase in pain of ≥20 mm was defined as a pain flare (29). Data were summarized for the first 14 days of the intervention as well as for the entire intervention period. |
| Motivation to perform the prescribed exercises, at baseline. | A self-developed questionnaire | The participants fulfilled a short questionnaire comprising three questions developed for this purpose, each with possible responses ordered in 4 levels on an ordinal scale. The questionnaire is available online as extended data in the protocol article (1, 14) |

| | | |
|---|---|---|
| Evaluation of the prescribed exercises, 10 weeks after surgery | A self-developed questionnaire | The participants fulfilled a short questionnaire comprising three questions developed for this purpose, each with possible responses ordered in 4 levels on an ordinal scale. The questionnaire is available online as extended data in the protocol article (1, 14) |
| Patient-perceived result after surgery, 10 weeks after surgery | A question phrased "How would you describe the result of your operation?" with response categories "Excellent", "Very good", "Good", "Fair", "Poor" (15) | The questions used for measuring patient-perceived result after surgery and change in hip problems have previously been used as anchor questions when establishing patient acceptable symptom state (PASS) and minimal clinically important improvement (MCII) cut-points for HOOS 1 year after THA (15). |
| Patient-perceived change in hip problems (from pre-surgery to 10 weeks after surgery), 10 weeks after surgery | A question phrased "Overall, how are the problems now in the hip on which you had surgery, compared to before your operation?" with the response categories "Much better", "A little better", "About the same", "A little worse", "Much worse" (15). | |

* HOOS: Hip disability and Osteoarthritis Outcome Score

### Appendix A - Supplements to the methods section

Appendix to: *Hip strengthening exercise dosage is not associated with clinical improvements after total hip arthroplasty – a prospective cohort study (the PHETHAS-1 study)*

**Supplementary description of data management**
Due to substantial differences between automatically software-generated and manually-counted number of repetitions, we manually counted every single repetition for all exercise sets. If manually determined number of repetitions was equal to the automatically generated number, the count was considered correct. If the numbers differed, another investigator also counted the repetitions. In case of disagreement, the two investigators visually inspected the data together until consensus was reached. In some cases, the heterogeneity of illustrated repetitions made counting very challenging. In these cases, the two investigators discussed the issue and labelled the counted number of repetitions with an interpretation level of either low (heterogeneity, but not really in doubt of the number of repetitions), medium (heterogeneity, but only a little doubt of the number of repetitions) or high (heterogeneity with some doubt of the number of repetitions (could be +/- a couple of repetitions)). In few exercise sets, the two investigators considered the counted number of repetitions more imprecise than what was suitable for the interpretation level "high". Here, the number of repetitions was not counted. Following the counting of repetitions, date and time of exercise sessions and number of repetitions were extracted.

Interpretation was used in about half of the participants. In most cases, interpretation levels were low and only occasionally used, but in three participants, a frequent and high level of interpretation was used (details in Appendix B). Furthermore, in one participant, data quality was too poor to calculate or count repetitions, and in further two cases, due to sensor failure, no data were obtained. Hence, the latter three cases were not included in the primary analysis. (Figure 1).

**Supplementary description of statistical methods used in the exploratory analyses and overview of handling of quantitative continuous and categorical data (e.g. grouping and transformation).**

***Exploratory analyses***
Association between performed exercise dose (variables: number of repetitions per week and number of exercise days per week) and several independent variables was investigated using univariate modelling. A linear regression model with fixed increase was used as first choice, but the approach was changed if model assumptions were not fulfilled (se details in 'handling of quantitative vriables'). Dependent Independent variables were: a) pain flares during the first two weeks of intervention, b) pain flares during the entire intervention period, c) HOOS_pain at baseline, d) self-efficacy at baseline, e) motivation to perform exercises, f) self-belief in compliance to perform exercises, g) belief in effect of exercises, h) satisfaction with rehabilitation exercise, i) mean upright time/day and j) mean number of steps/day.
Association between physical activity (variables: mean upright time/day and mean number of steps/day) and several independent variables was investigated using a univariate linear regression model with fixed increase. Independent variables were: a) pain flares during the first two weeks of intervention, b) HOOS_pain at baseline, c) self-efficacy at baseline, d) motivation to perform exercises and e) self-belief in compliance to perform exercises.

In the analysis of patient-perceived result of surgery, the change in gait speed was presented in medians with 1st and 3rd quartiles for each response category, as well as for the subgroup of participants, who answered “excellent”, “very good” or “good”. This subgroup, was considered to have achieved a hip-specific acceptable symptom state (PASS). Similarly, the change in each HOOS subscale was presented for the same categories, but with data being presented as mean scores with 95% CIs. In addition, the percentage of participants in each response category is illustrated in bar charts distributed on exercise quartiles. Furthermore, using the scores at 10week, HOOS cut points for PASS were estimated. Cut points were presented in both median change (as data were not normally distributed) and mean change, to allow for comparison with previous estimates (15).

In the analysis of patient-perceived change in hip problems, the change in gait speed was presented in medians with 1st and 3rd quartiles for each response category. Similarly, the change in each HOOS subscale was presented for the same categories, but with data being presented as mean scores with 95% CIs. In addition, the percentage of participants in each response category is illustrated in bar charts distributed on exercise quartiles. Finally, it was planned to estimate cut points for MCII, but due six observations only in the response category "a little better", it did not make sense to perform this analysis.

***Handling of quantitative variables in the analyses***

Continuous variables

- In general, continuos variables are analysed as they are.
- In some of the primary and secondary analyses, the population was divided in four groups based on quartiles of performed exercise dose (mean number of repetitions per week).
- In the secondary analysis, when testing association between the independent variables and gait speed at 10 weeks, a logarithmic transformation of outcome was neded to fulfill the presumtions for the pre-defined multiple linear regression model.
- In the exploratory analysis, when testing of association between the continous independent variables and exercise dose, a linear regression model was first choice. If the presumptions for the model were not fullfilled, logarithmic transformation of continous outcome variables or insertion of polynomiums in the model were tried, followed by transforming continous variables to categorical.
    - This approach led to grouping of the following continous variables: HOOS_pain, self-efficacy, upright time per day and step per day were categorised in quartiles as a proxy for linear asociation. Pain flare was categorised in two ways based on a combination of data distribution and clinical reasoning. In one analysis, the categories were: a) no pain flares, b) one pain flare and 3) more than one pain flare. In the other analysis, the categories were: a) no pain flares and b) at least one pain flare.
- After grouping, if presumptions for linear regression were still not fulfilled, comparison of data distribution between categories were made using Kruskal Wallis test (as presumptions for ANOVA were not fulfilled).
- Based on the above approach, the association is tested differently among the independent continous variables.

Categorical variables

- In general, categorical variables are analysed using their original categories.
- In the exploratory analysis, when testing of association between the categorical independent variables and exercise dose, some variable categories were collapsed into two categories

only to explore more simple associations. The choice of how to collapse categories was made based on both clinical reasoning and data distribution.
    - Motivation to perform exercises was divided in the categories: a) very much and b) less than very much. Self-belief in compliance to exercise was divided in the categories: a) very certain and b) less than very certain. Satisfaction with rehabilitation exercises was divided in the categories: a) satisfied or very satisfied and b) unsatisfied or very unsatisfied.
- Based on the above approach, the association is tested differently among the independent variables.
