## Appendix B for "Hip strengthening exercise dosage is not associated with clinical improvements after total hip arthroplasty – a prospective cohort study (the PHETHAS-1 study)"

### Supplementary description on participant characteristics

**Table B1.** Summary statistics on prosthesis type and previous joint arthroplasty

| | | |
|---|---|---|
| **Supplementary descriptive variables** | **Prosthesis type**<br>*number (percentage)* | |
| | Fixation, n=67 | |
| | - Uncemented | 67 (100) |
| | Stem, n=91 | |
| | - Corail | 53 (58) |
| | - Avenir | 27 (30) |
| | - Spotorno | 10 (11) |
| | - Wagner cone | 1 (1) |
| | Cup, n=51 | |
| | - Pinnacle | 38 (75) |
| | - Trident | 10 (20) |
| | - Gripton | 3 (6) |
| | Previous joint arthroplasty, n=91<br>*number (percentage)* | |
| | - None | 55 (60) |
| | - No THA (TKA unknown) | 4 (4) |
| | - THA | 28 (31) |
| | - TKA | 3 (3) |
| | - THA and TKA | 1 (1) |

### Supplementary description on exercise dose

The length of intervention varied from 36 to 65 days with a mean of 50 [CI: 49-51] days. Fifty percent of the participants had an intervention period between 6.7 and 7.4 weeks and 85% had an intervention lasting 7 weeks +/- one week. During the intervention, 85 % of the participants used the sensor technology every time or most of the time during exercising. Summary statistics on performed exercise dose is presented in Table B2.

**Table B2.** Summary statistics on performed exercise dose based on data from sensor technology attached to participants' elastic band

| | | |
|---|---|---|
| **Performed exercise dose** | **Number of repetitions**, n=91<br>*median (1st and 3rd quartile)* | |
| | - total | 2634 (1440; 3922) |
| | - per week | 339 (209; 549) |
| | **Exercise sessions** (days), n=92<br>*median (1st and 3rd quartile)* | |
| | - total | 19 (14.5; 23) |
| | - per week | 2.7 (2.0; 3.2) |

*More than one session per day*

In total, the population performed strengthening exercises in 1679 days. In 100 of these, exercises were performed more than once that day distributed on 18 participants. Among these 18, 3 participants performed exercises more than once per day on at least 20 of their exercise days, one participant in 8 days, while the remaining 14 participants performed exercises more than once per day three days or less.

*Interpretation used*
One participant's data were too complex to be able to calculate/count number of repetitions. Among the remaining 91 participants 56 participants had at least one set of repetitions, where some kind of interpretation in calculating/counting reps was used. In 40 cases, numbers of days with interpretation were below 4, in 13 cases the numbers were 4-9 and in the last three cases, interpretation was used at least 16 times.
Among the 56 participants, where interpretation was used, the proportion of days with interpretation used varied between 0.03 and 0.95. Median proportion was 0.125 (IQR [0.07; 0.23]). In three cases, the proportion was above 0.83.
Among the 56 participants, number of times interpretation were used varied between one and 68. 45 participants had 10 or less interpretations, six persons varied from 11-28, two were in the forties, while three had more than 60 times, where interpretation was used.
The level of interpretation was registered as low, moderate or high. When using weight of severity of interpretation (low – weight 1, moderate – weight 2, high – weight three), the levels of interpretations varied between one and 171. 40 participants ranged from 1-10, 10 participants from 12-35, 3 participants between 56-87 and three participants were above 100. Regardless of which method used for evaluating degree of interpretation, the same three participants were identified as the ones with the highest levels.

### Supplementary description on motivation for home-based rehabilitation exercise, evaluation of exercise and patient-perceived result of surgery

**Table B3.** Summary statistics on items regarding motivation for home-based rehabilitation exercise measured at baseline (3 weeks)

| **Motivation** | **Motivation to perform exercises**, n=93<br>*number (percentage)* | |
|---|---|---|
| | - Very much | 81 (87) |
| | - To some degree | 11 (12) |
| | - A little | 1 (1) |
| | - Not at all | 0 (0) |
| | - Don't know | 0 (0) |
| | **Belief in effect of exercises**, n=93<br>*number (percentage)* | |
| | - Very much | 88 (95) |
| | - To some degree | 5 (5) |
| | - A little | 0 (0) |
| | - Not at all | 0 (0) |
| | - Don't know | 0 (0) |
| | **Self-belief in compliance to exercise**, n=93<br>*number (percentage)* | |
| | - Very certain | 50 (54) |
| | - Almost certain | 42 (45) |
| | - A little uncertain | 1 (1) |
| | - Very uncertain | 0 (0) |
| | - Don't know | 0 (0) |

**Table B4**. Summary statistics on items evaluating prescribed exercise and describing patient-perceived result measured at 10-week follow-up.

| | | |
|---|---|---|
| **Evaluation of prescribed exercises** | **Satisfaction with rehabilitation exercises**, n=92 *number (percentage)* | |
| | - Very satisfied | 65 (71) |
| | - Satisfied | 21 (23) |
| | - Unsatisfied | 4 (4) |
| | - Very unsatisfied | 1 (1) |
| | - Don't know | 1 (1) |
| | **Study-related change of exercise adherence**, n=92 *number (percentage)* | |
| | - Exercised more | 70 (76) |
| | - Exercised the same | 21 (23) |
| | - Exercised less | 0 (0) |
| | - Don't know | 1 (1) |
| | **Compliance with BandCizer**, n=93 *number (percentage)* | |
| | - Every time | 68 (73) |
| | - Most of the time | 11 (12) |
| | - About half the time | 7 (8) |
| | - A few times | 6 (6) |
| | - Never | 1 (1) |
| **Patient-perceived result** | **Rating of perceived result**, n=89 *number (percentage)* | |
| | - Excellent | 58 (65) |
| | - Very good | 23 (26) |
| | - Good | 4 (4) |
| | - Fair | 2 (2) |
| | - Poor | 2 (2) |
| | **Change in hip problems**, n=89 *number (percentage)* | |
| | - Much better | 78 (88) |
| | - A little better | 6 (7) |
| | - About the same | 4 (4) |
| | - A little worse | 1 (1) |
| | - Much worse | 0 (0) |

**APPENDIX B - Supplementary descriptive results**
Appendix to: *Hip strengthening exercise dosage is not associated with clinical improvements after total hip arthroplasty – a prospective cohort study (the PHETHAS-1 study)*

**Figure B1.** Patient-perceived result of surgery across quartile groups of performed number of repetitions per week (exercise dosages)

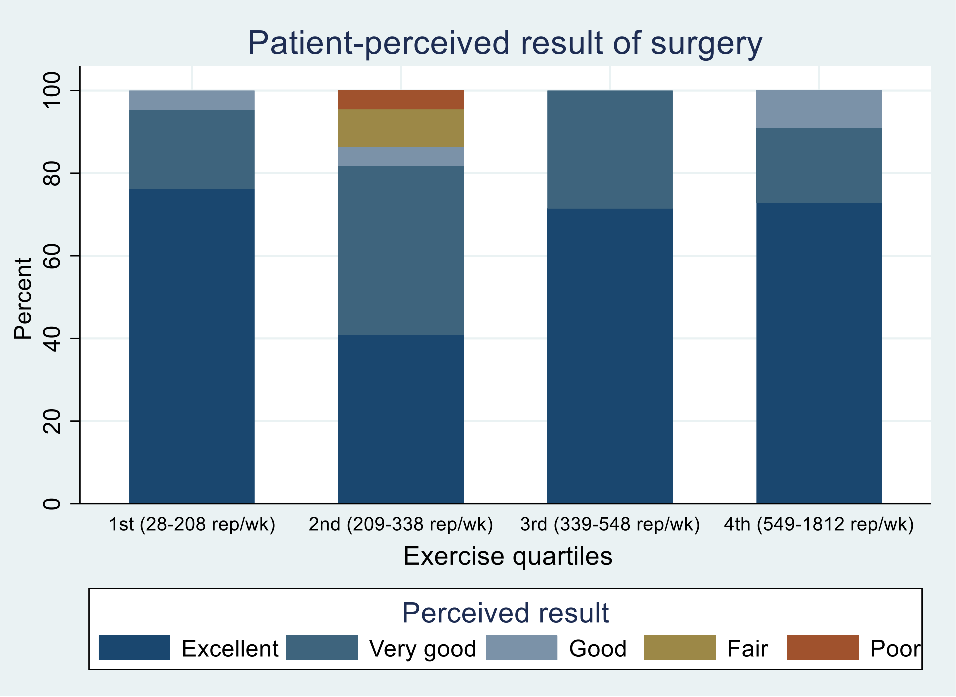


**Supplementary description on exercise-related pain and adverse events**

**Table B5.** Summary statistics on exercise-related pain measured as change in pain at rest before exercise and pain at rest after exercise. Pain is measured in mm using visual analogue scale (VAS). Pain flare is defined as an increase of at least 20 mm.

| | | |
|---|---|---|
| **Pain** | **Change in pain at rest (mm) per exercise session**, n=93<br>*Median (IQR)* | 1.45 (0.2; 3.72) |
| | **Pain flares during the first 14 days**<br>(n=93, sessions: n=554)<br>- Sessions with pain flare<br>*number (percentage)*<br>- Participants with at least one pain flare<br>*number (percentage)* | 36 (6)<br><br>18 (19) |
| | **Pain flares during the entire period**<br>(n=93, sessions: n=1583)<br>- Sessions with pain flare<br>*number (percentage)*<br>- Participants with at least one pain flare<br>*number (percentage)* | 57 (4)<br><br>21 (23) |

**Supplementary description on pain**
According to diaries, the participants have performed strengthening exercises in a varying number of days ranging from 2 to 46. In total, strengthening exercise was performed in 1853 days. Median number of days per participant was 21(IQR: 16-25). During the first 14 days, a total of 612 exercise sessions were performed.
Pain-change is based on data from 1583 exercise days in 93 of 94 participants. For each participant, the average pain-change is used to calculate average pain-change in the

population. In 27 participants, pain-change data is available for all exercises sessions. Across the population, the median proportion of pain-change data available for the exercise session is 92% (IQR: 80-100)
Pain flare is registered in 57 exercise sessions distributed on 21 participants. Especially one participant had a large number of flare-ups (15 of 24 sessions with pain-change data, 25 sessions registered).
During the first 14 days, pain change data were available in 554 sessions. Pain flares occurred 36 times distributed on 18 participants. One participant had 7 pain flares, 3 participants each had 4 pain flares, while 14 participants experienced one or two pain flares.

**Table B6.** Summary statistics on serious and non-serious adverse events occurring between surgery and 10-week follow-up.

| **Adverse events** | **Serious adverse events**, n=89 | |
|---|---|---|
| | *Number (percentage)* | |
| | - None | 87 (98) |
| | - Hip dislocation | 0 (0) |
| | - Infection (deep) | 0 (0) |
| | - Fracture | 0 (0) |
| | - Acute myocardial infarction | 0 (0) |
| | - Deep veinthrombosis | 0 (0) |
| | - Readmission | 2 (1)* |
| | - Other | 0 (0) |
| | **Non-serious adverse events**, n=87† | |
| | *Number (percentage)* | |
| | - None | 58 (67) |
| | - Wound seepage/bleeding | 4 (5) |
| | - Wound infection | 4 (5)‡ |
| | - Other§ | 21 (24) |

* Readmission caused by bleeding or wound seepage. Further details not reported due to risk of indirect identification.
† The subpopulation of 87 participants, where no serious advents occurred
‡ In three cases wound seepage/bleeding were also registered
§ Complications/problems, which have caused the participant to contact a health care person: vasovagal coincidence (n=1), superficial flebitis (n=1), haematoma/oedema (n=4), itching (n=2), pain (n=10) (no further details due to risk of indirect identification), stiffness when initiating movement (n=2), hypotension/low haemoglobolin (n=1), clicking from the hip (n=1), concern of wound infection – not confirmed (n=1), other (non-hip) fracture (n=1), bullae from wound dressing (n=1)
