## Appendix C for "Hip strengthening exercise dosage is not associated with clinical improvements after total hip arthroplasty – a prospective cohort study (the PHETHAS-1 study)"

**Appendix C – Supplementary results on secondary and exploratory analyses**
Appendix to: *Hip strengthening exercise dosage is not associated with clinical improvements after total hip arthroplasty – a prospective cohort study (the PHETHAS-1 study)*

### Secondary analysis

**Figure C1.** Box plots of the 3 (baseline) to 10-week (follow-up) median change in symptoms measured by HOOS, for quartile groups of performed number of repetitions per week (exercise dosages).

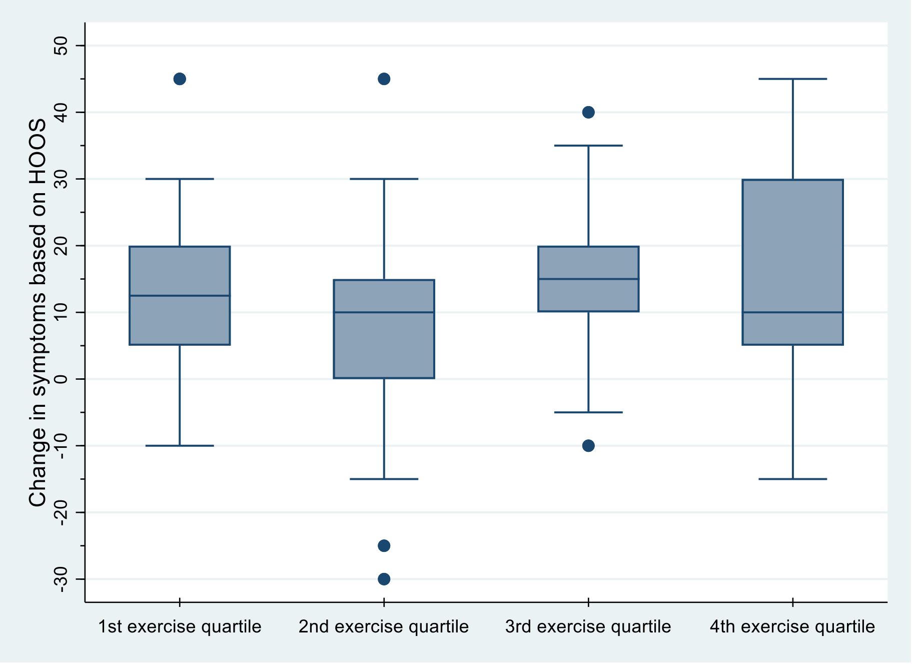


**Figure C2.** Box plots of the 3 (baseline) to 10-week (follow-up) median change in quality of life measured by HOOS, for quartile groups of performed number of repetitions per week (exercise dosages)

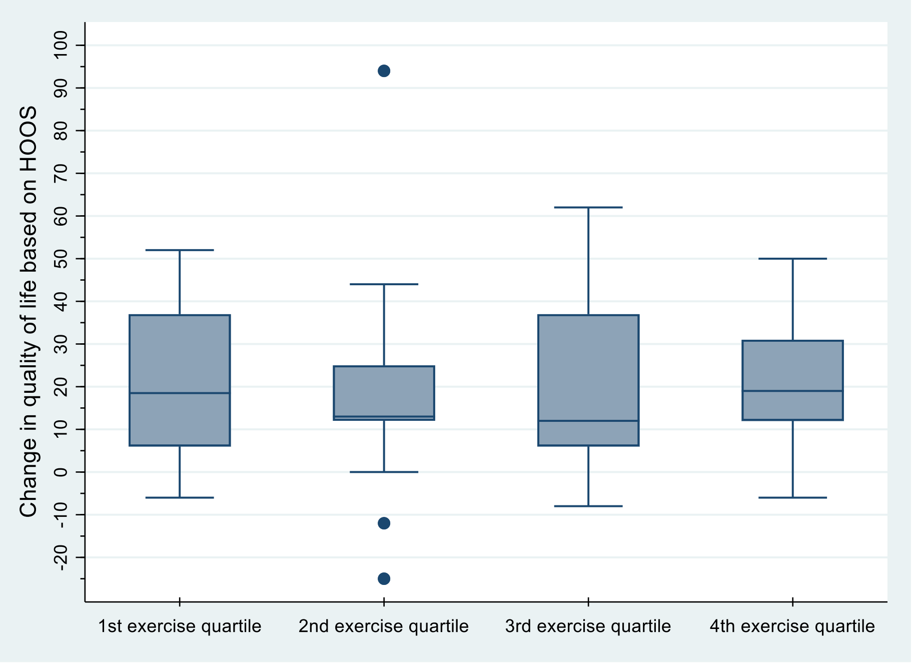

**Figure C3.** Box plots of the 3 (baseline) to 10-week (follow-up) median change in isometric hip abduction strength (Nm/kg), for quartile groups of performed number of repetitions per week (exercise dosages).

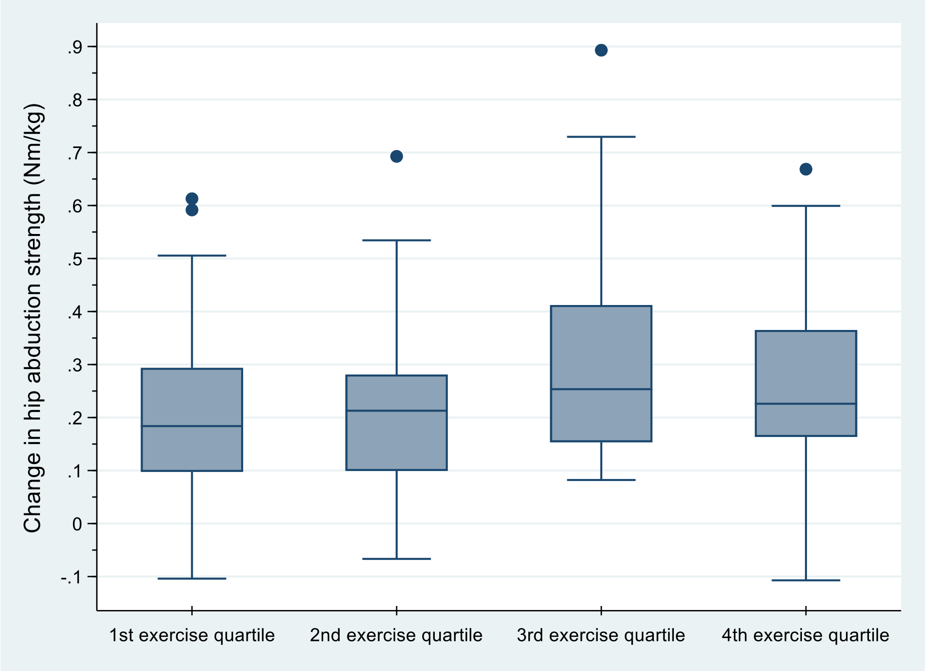


**Figure C4.** Box plots of the 3 (baseline) to 10-week (follow-up) median change in isometric hip flexion strength, for quartile groups of performed number of repetitions per week (exercise dosages).

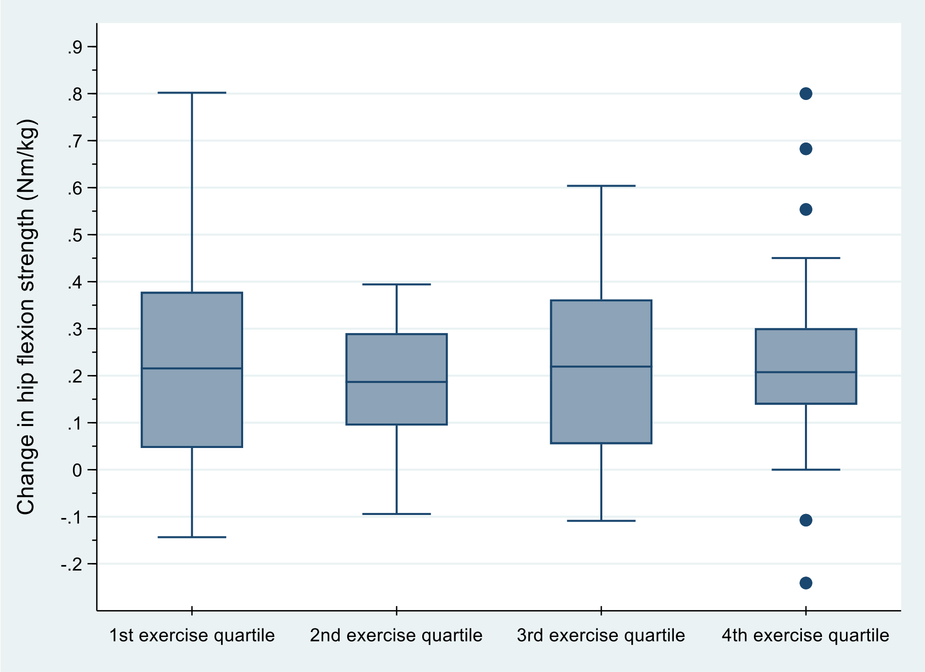

**Appendix C – Supplementary results on secondary and exploratory analyses**
Appendix to: *Hip strengthening exercise dosage is not associated with clinical improvements after total hip arthroplasty – a prospective cohort study (the PHETHAS-1 study)*

**Table C1.** Change in outcomes from baseline (3 weeks after surgery) to follow-up (10 weeks after surgery) distributed on quartiles of performed exercise dose.

| | 1st exercise quartile (28-208 reps/week) n=22 | 2nd exercise quartile (209-338 reps/week) n=23 | 3rd exercise quartile (339-548 reps/week) n=23 | 4th exercise quartile (549-1812 reps/week) n=23 |
|---|---|---|---|---|
| **Gait speed,** m/sec *mean (95% CI)* | 0.26 (0.18; 0.33) | 0.32 (0.25; 0.39) | 0.37 (0.28; 0.47) | 0.37 (0.30; 0.45) |
| **HOOS_adl** *mean (95% CI)* | 13 (8; 19) | 10 (5; 15) | 16 (11; 21) | 16 (10; 22) |
| **HOOS_pain** *mean (95% CI)* | 14 (8; 19) | 10 (4; 15)* | 16 (9; 22) | 14 (8; 20) |
| **HOOS_sym** *mean (95% CI)* | 14 (7; 20) | 6 (-1; 14)* | 15 (10; 20) | 14 (8; 21) |
| **HOOS_qol** *mean (95% CI)* | 19 (11; 27) | 18 (8; 29)† | 20 (12;28) | 23 (17; 29) |
| **30s chair-stand** *mean (95% CI)* | 3.9 (2.0; 5.8) § | 4.6 (2.6; 6.5) § | 6.0 (4.4; 7.5) | 5.0 (3.8; 6.1)‡ |
| **Hip abduction strength,** Nm/kg *mean (95% CI)* | 0.22 (0.13; 0.30) | 0.22 (0.13; 0.30)‡ | 0.32 (0.23; 0.41) | 0.26 (0.18; 0.33) |
| **Hip flexion strength,** Nm/kg *mean (95% CI)* | 0.24 (0.14; 0.34) | 0.18 (0.12; 0.24)* | 0.22 (0.14; 0.31)* | 0.24 (0.14; 0.34) |

* n=22, † n=21, ‡ n=20, § n=18

**Table C2.** Association between gait speed at 10 weeks follow up and the independent variables: performed exercise dose (mean number of repetitions/week), self-efficacy at baseline, 24-hour physical activity (Mean upright time/day and mean number of steps/day), and gait speed at baseline. The association is presented as relative increase.

| **Independent variables** | **Coefficient (95% CI)** |
|---|---|
| Mean number of repetitions/week (in hundreds) | 1.007 (0.998; 1.017) |
| Self-efficacy at baseline | 1.048 ( 0.995; 1.104) |
| Mean upright time/day | 0.981 (0.964; 0.998) |
| Mean number of steps/day | 1.000 (1.000; 1.000) |
| Gait speed at baseline | 1.005 (1.004; 1.005) |

**Appendix C – Supplementary results on secondary and exploratory analyses**
Appendix to: *Hip strengthening exercise dosage is not associated with clinical improvements after total hip arthroplasty – a prospective cohort study (the PHETHAS-1 study)*

### Exploratory analyses

#### Factors associated with exercise dose

Based on the statistical approach thorougly described in Appendix A, the association is tested differently among the independent variables.
For HOOS_pain at 3 weeks and Self-efficacy at 3 weeks, the association could be presented as a polynomial equation along with presentation of a graphical prediction line. To show further details on distribution of data, analyses based on quartiles – along with Kruskall-Wallis test – is also presented.

**Table C3.**
Factors associated with exercise dose in terms of repetitions per week and exercise days per week.

| Dependent variable / Independent variable | **Repetitions per week**<br>Median (1st-3rd quartile) | p-values (Kruskall-Wallis test) * | **Exercise days per week**<br>Median (1st-3rd quartile) | p-values (Kruskall-Wallis test) * |
|---|---|---|---|---|
| **Number of pain flares (week 0-2)**, n=91 | | | | |
| - No pain flares, n=73† | 339 (208-561) | p=0.82 | 2.7 (2-3.2) | p=0.97 |
| - One pain flare, n=11 | 462 (246-521) | | 3 (2.0-3.2) | |
| - Two or more pain flares, n=7 | 257 (144-500) | | 2.6 (1.4-3.3) | |
| - No pain flares, n=73† | 339 (208-561) | p=0.76 | 2.7 (2-3.2) | p=0.83 |
| - At least one pain flare, n=18 | 383.5 (246-500) | | 2.8 (2.0-3.2) | |
| **Number of pain flares (entire period)**, n=91 | | | | |
| - No pain flares, n=71† | 370 (182-572) | p=0.66 | 2.7 (2-3.2) | p=0.85 |
| - One pain flare, n=10 | 391 (302-521) | | 3.0 (2.0-3.2) | |
| - Two or more pain flares, n=10 | 254.5 (209-470) | | 2.6 (2.4-3.2) | |
| - No pain flares, n=71† | 370 (182-572) | p=0.59 | 2.7 (2-3.2) | p=0.78 |
| - At least one pain flare, n=20 | 312.5 (234-485) | | 2.9 (2.2-3.2) | |
| **HOOS_pain at 3 week**, n=90† | Association is visualized in Figure 12 | p=0.11‡ | | |
| - 1st quartile (40-65), n=26† | | | 2.6 (2.0-3.1) | p=0.43 |
| - 2nd quartile (68-75), n=21 | | | 3.1 (2.4-3.3) | |
| - 3rd quartile (78-88), n=21 | | | 2.8 (2.2-3.4) | |
| - 4th quartile (90-100), n=22 | | | 2.7 (1.6-3.1) | |
| **Self-efficacy at 3 week**, n=75 | Association is visualized in Figure 13 | p= 0.38§ | | |
| - 1st quartile (1-2.9), n=18 | | | 3.1 (2-3.3) | p=0.28 |
| - 2nd quartile (3-3.4), n=24 | | | 2.8 (2.2-3.4) | |
| - 3rd quartile (3.5-3.8), n=24† | | | 2.6 (2.0-3.1) | |
| - 4th quartile (3.9-4), n=19 | | | 2.6 (1.7-3) | |
| **Motivation to perform exercises** | | | | |
| - Very much, n=78† | 373 (212-550) | | 2.8 (2.1-3.2) | |
| - To some degree, n=11 | 305 (144-492) | p=0.31 | 2.7 (1.4-3.3) | |
| - A little, n=1 | 99 (99-99) | | 0.3 (0.3-0.3) | p=0.23 |

| | | | | |
|---|---|---|---|---|
| - Not at all, n=0<br>- Don't know, n=0 | NA<br>NA | | NA<br>NA | |
| - Very much, n=78†<br>- Less than very much, n=12 | 373 (212-550)<br>275.5 (121.5-477.5) | p=0.28 | 2.8 (2.1-3.2)<br>2.7 (1.3-3.2) | p=0.64 |
| **Belief in effect of exercises**<br>- Very much, n=85†<br>- To some degree, n=5<br>- A little, n=0<br>- Not at all, n=0<br>- Don't know, n=0 | <br>376 (209-549)<br>257 (222-305)<br>NA<br>NA<br>NA | p=0.40 | <br>2.8 (2.0-3.2)<br>2.7 (2.6-3.1)<br>NA<br>NA<br>NA | p=0.98 |
| **Self-belief in compliance to exercise**<br>- Very certain, n=49<br>- Almost certain, n=40†<br>- A little uncertain, n=1<br>- Very uncertain, n=0<br>- Don't know, n=0 | <br>418 (175-550)<br>323 (222-510.5)<br>222 (222-222)<br>NA<br>NA | p=0.72 | <br>2.9 (2.1-3.2)<br>2.6 (2-3.2)<br>2.7 (2.7-2.7)<br>NA<br>NA | p=0.71 |
| - Very certain, n=49<br>- Less than very certain, n=41† | 418 (175-550)<br>308 (222-500) | p=0.69 | 2.9 (2.1-3.2)<br>2.6 (2-3.2) | p=0.41 |
| **Satisfaction with rehabilitation exercises**<br>- Very satisfied, n=63<br>- Satisfied, n=21<br>- Unsatisfied, n=3†<br>- Very unsatisfied, n=1<br>- Don't know, n=1 | <br>418 (208-589)<br>320 (222-539)<br>214 (52-252)<br>246 (246-246)<br>68 (68-68) | p=0.19 | <br>2.8 (2.1-3-2)<br>3 (2.1-3.4)<br>1.9 (0.9-2.3)<br>2.0 (2.0-2.0)<br>0.6 (0.6-0.6) | p=0.11 |
| - Satisfied or very satisfied, n=84<br>- Unsatisfied or very unsatisfied, n=4† | 230 (133-249)<br>379.5 (210.5-555.5) | p=0.07 | 2.8 (2.1-3.2)<br>2.0 (1.5-2.3) | p=0.03 |
| **Upright time per day** (hours), n=78 | 44 (3; 84), p=0.04 ‖ | | | |
| - 1st quartile (2.58-4.39), n=20<br>- 2nd quartile (4.41-5.6), n=20<br>- 3rd quartile (5.69-6.56), n=20<br>- 4th quartile (6.58-9.34), n=19 | | | 2.8 (2.0-3.2)<br>2.6 (2.1-3.1)<br>2.5 (1.6-3.3)<br>2.9 (2.3-3.2) | P=0.78 |
| **Steps per day** (in 1000), n=78 | 36 (13; 58), p=0.003 ‖ | | | |
| - 1st quartile (1748-4744), n=20<br>- 2nd quartile (4887-6019), n=20<br>- 3rd quartile (6346-8239), n=20<br>- 4th quartile (8907-14188), n=19 | | | 2.7 (2.3-3.1)<br>2.3 (1.8-3)<br>2.8 (1.9-3.1)<br>3.1 (2.3-3.4) | p=0.28 |

* unless otherwise stated
† plus one extra observation i analysis of association between exercise days and independent variable
‡ a non-linear regression model (third-grade polynomium) was used.
§ a non-linear regression model (fourth-grade polynomium) was used.
‖ A linear regression model was used

**Figure C5.** Association between HOOS_pain at baseline and performed number of repetitions per week.

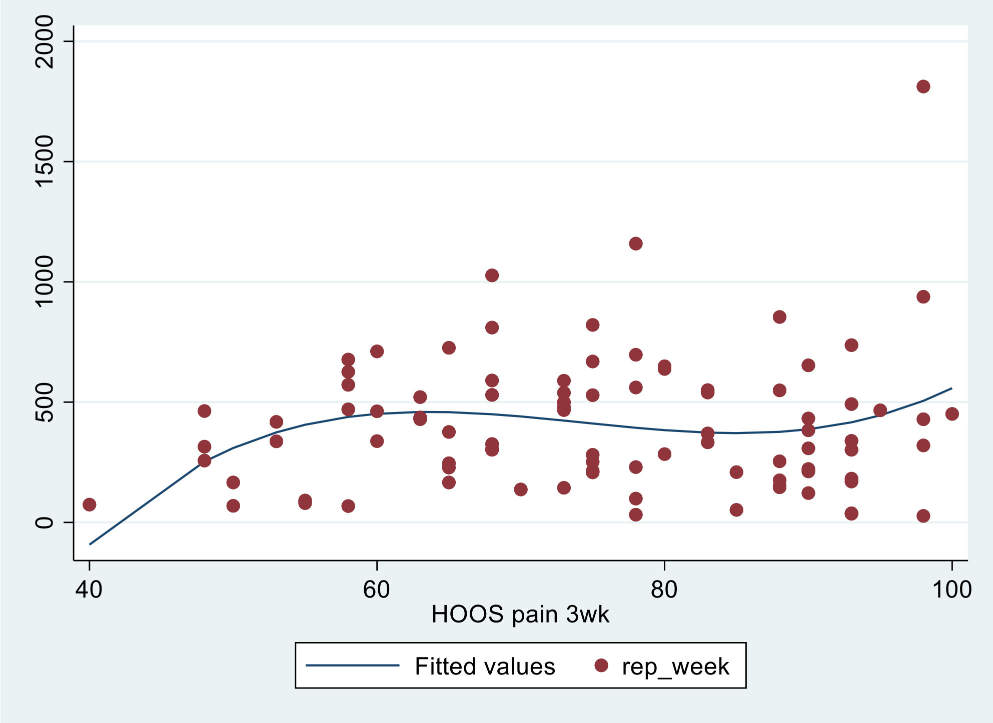


**Figure C6.** Association between mean self-efficacy at baseline and performed number of repetitions per week.

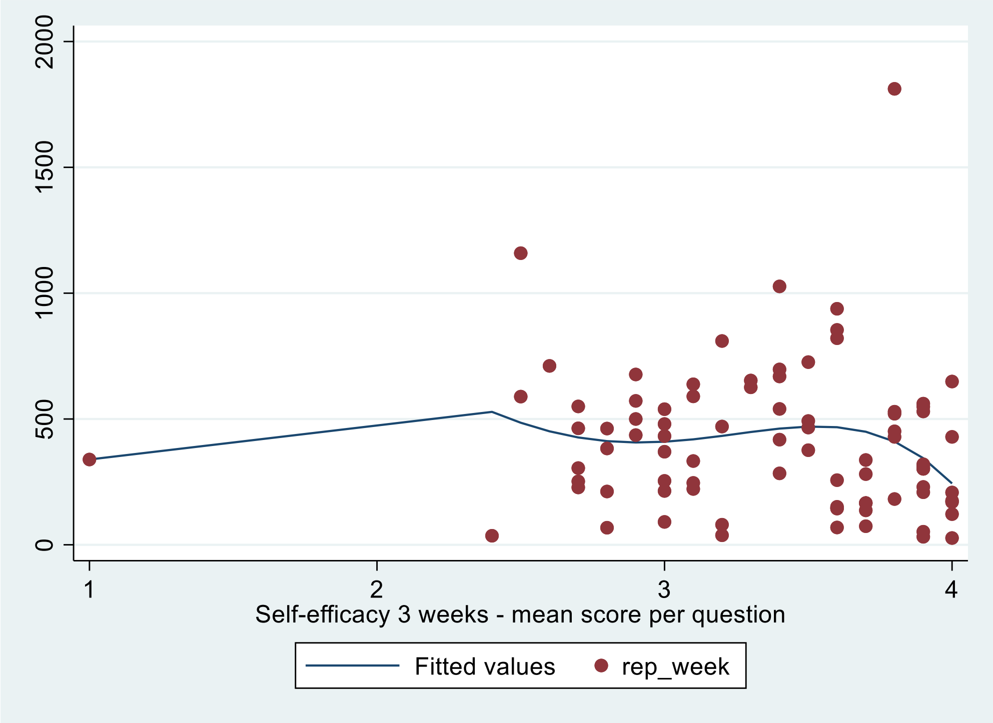

### Appendix C – Supplementary results on secondary and exploratory analyses

Appendix to: *Hip strengthening exercise dosage is not associated with clinical improvements after total hip arthroplasty – a prospective cohort study (the PHETHAS-1 study)*

#### Factors associated with physical activity

**Table C4.** Factors associated with physical activity in terms of upright time per day and steps per day. Association is determined by univariate linear regression models

| Dependent variable / Independent variable | **Upright time/day** (hours) | **Step/day** (numbers) |
|---|---|---|
| **Number of pain flares (week 0-2)**, n=81 | -0.14 (-0.55; 0.26) | -118 (-828; 592) |
| **HOOS_pain at 3 week**, n=80 | -0.01 (-0.03; 0.01) | -41 (-80; -2)* |
| **Self-efficacy at 3 week**, n=75 | -0.30 (-0.98; 0.38) | 617 (-595; 1829) |
| Motivation to perform exercises | | |
| - Very much, n=70 | *Reference* | *Reference* |
| - To some degree, n=9 | -0.55 (-1.64; 0.53) | -763 (-2695; 1169) |
| - A little, n=1 | -2.37 (-5.45; 0.71) | 164 (-5330; 5658) |
| - Not at all, n=0 | NA | NA |
| - Don't know, n=0 | NA | NA |
| Very much versus lesser degree of motivation (very much is reference value) | -0.73 (-1.77; 0.30) | -670 (-2503; 1163) |
| Self-belief in compliance to exercising | | |
| - Very certain, n=43 | *Reference* | *Reference* |
| - Almost certain, n=37 | 0.37 (-0.32; 1.06) | -525 (-1739; 690) |
| - A little uncertain, n=0 | NA | NA |
| - Very uncertain, n=0 | NA | NA |
| - Don't know, n=0 | NA | NA |

*p=0.04

#### Patient-perceived change in hip symptoms and MCII

**Table C5.** Change from baseline(3 weeks) to 10-week follow-up in gait speed, HOOS_adl, HOOS_symp, HOOS_pain and HOOS_qol, distributed on level of patient-perceived change in hip symptoms

| | **Change in HOOS_adl** mean (95% CI) | **Change in HOOS_pain** mean (95% CI) | **Change in HOOS_symp** mean (95% CI) | **Change in HOOS_qol** mean (95% CI) | **Change in gait speed** *(m/sec)* median [1st-3rd quartile] |
|---|---|---|---|---|---|
| **Much better,** n=77 | 14 (11; 17)* | 14 (11; 17) | 14 (11; 16) | 22 (19; 27)* | 0.32 [0.21-0.42]† |
| **A little better,** n=6 | 15 (-5; 36) | 13 (-5; 30) | 18 (-6; 41) | 10 (-4; 24) | 0.36 [0.30-0.41] |
| **About the same,** n=4 | 9 (-12; 30) | -4 (-23; 15) | -4 (-32; 25) | 3 (-31; 37) | 0.20 [0.18-0.23] |
| **A little worse,** n=1 | 6 (NA) | 12 (NA) | -25 (NA) | 0 (NA) | 0.36 [NA] |
| **Much worse,** n=0 | NA | NA | NA | NA | NA |

* n=76, † n=78

**Figure C7.** Distribution of level of patient-perceived change in hip symptoms presented for each quartile of performed number of repetitions per week

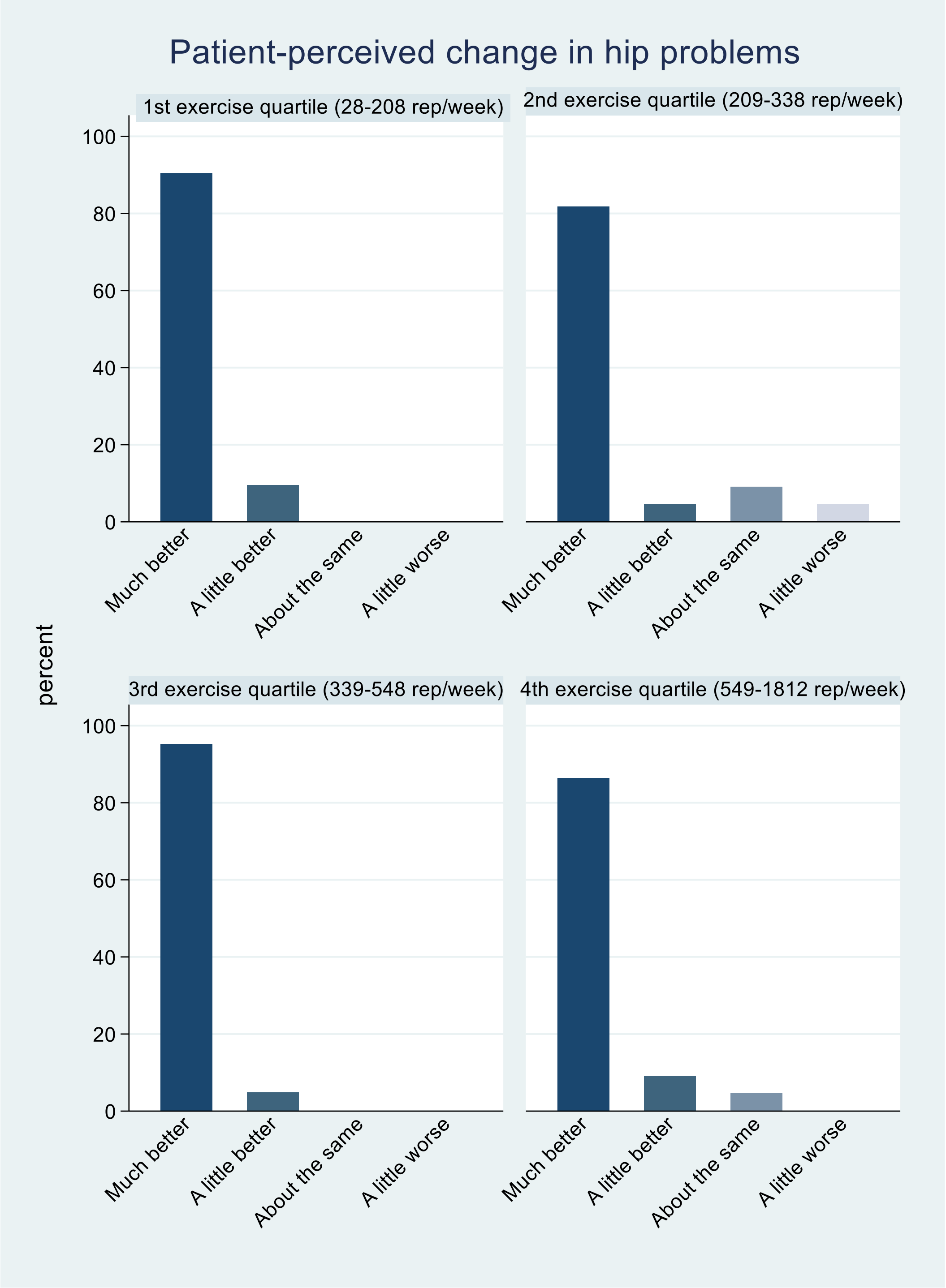


**MCII**
Due to only 6 observations reporting change in hip symptoms to be "a little better", the estimate of MCII was very imprecise. Results can be seen in tableC4.

### Patient-perceived result of surgery

**Table C6.** Change from baseline to 10-week follow-up in gait speed, HOOS_adl, HOOS_symp, HOOS_pain and HOOS_qol, distributed on level of patient-perceived result of surgery

| | **Change in HOOS_adl** mean (95% CI) | **Change in HOOS_pain** mean (95% CI) | **Change in HOOS_symp** mean (95% CI) | **Change in HOOS_qol** mean (95% CI) | **Change in gait speed** *(m/sec)* median [1st-3rd quartile] |
|---|---|---|---|---|---|
| **Excellent** | n=56 14 (11; 18) | n=57 14 (11; 18) | n=57 13 (10; 16) | n=57 22 (17; 27) | n=58 0.32 [0.20-0.43] |
| **Very good** | n=23 13 (8; 17) | n=23 13 (8; 18) | n=23 15 (10; 20) | n=23 20 (11; 30) | n=23 0.34 [0.27-0.40] |
| **Good** | n=4 22 (7; 37) | n=4 17 (-7; 41) | n=4 15 (-13; 43) | n=4 22 (9; 35) | n=4 0.34 [0.25-0.40] |
| **Fair** | n=2 [6-10]* | n=2 [-10-12]* | n=2 [-25-10]* | n=2 [0-12]* | n=2 [0.25-0.36]* |
| **Poor** | n=2 [-10-18]* | n=2 [-17-10]* | n=2 [-30-5]* | n=2 [-25-0]* | n=2 [0.16-0.21]* |
| **Acceptable†** | n=83 14 (11; 17) | n=84 14 (11;17) | n=84 14 (11;16) | n=83 21 (17; 25) | 85 0.33 [0.21-0.42] |
| **Not acceptable‡** | n=4 6 (-13; 25) | n=4 -1 (-24; 22) | n=4 -10 (-42; 22) | n=4 -3 (-28; 22) | 4 0.23 [0.18-0.31] |

* Range
† Comprises participants reporting an excellent, very good or good result of the operation
‡ Comprises participants reporting a fair or poor result of the operation

**PASS**

**Table C7.** Mean and median scores for HOOS subscales at 10 week follow-up for participants reporting a good, very good or excellent result of the operation.

| | **Mean (95% CI)** | **Median [1st-3rd quartile)** |
|---|---|---|
| **HOOS_adl**, n=84 | 89 (87; 91) | 91 [87.5-95.5] |
| **HOOS_symptoms**, n=85 | 84 (81; 86) | 85 [75-90] |
| **HOOS_pain**, n=85 | 90 (88; 93) | 93 [85-98] |
| **HOOS_qol,** n=84 | 78 (74; 81) | 75 [69-100] |
